# Distinct Systemic and Salivary Cytokine Profiles in Children with Perinatal HIV Exposure and Infection

**DOI:** 10.64898/2026.11.24.26368905

**Authors:** Aris G. Ilada, Ibukun John Abulude, Sukhwinder Singh, Esosa Osagie, Paul Akhigbe, Anil Kumar, Ozoemene Obuekwe, Patricia Bocarsly, Vincent P. Richards, Modupe Coker

## Abstract

**Background:** Despite antiretroviral therapy (ART), children living with HIV (HI) experience a disproportionate burden of oral diseases, including dental caries. It remains unclear whether this increased susceptibility is associated with altered local expression of inflammatory cytokines, and whether systemic inflammatory profiles reflect corresponding immune signals in saliva.

**Methods:** We analyzed 10 plasma and 7 salivary cytokines in matched plasma and unstimulated saliva samples from 467 Nigerian children aged 4–11 years: HI (n = 144), HEU (n = 161), and HUU (n = 162). Correlation analyses assessed plasma-saliva concordance, PERMANOVA evaluated overall cytokine profiles, multivariable linear regression examined individual cytokines, and network density analysis quantified connectivity.

**Results:** Cytokines correlated more strongly within than between compartments. HIV exposure and infection status were associated with overall cytokine profiles in plasma (R² = 0.008, p = 0.038) and saliva (R² = 0.017, p < 0.001), and pairwise differences between comparison groups were significant only in saliva. HI children had higher plasma IL-5 and MCP-1 and higher salivary IL-6 and MCP-1 than HEU and HUU children. Higher CD4⁺ T-cell count was associated with lower plasma IL-6 and MCP-1 and lower salivary IL-6. Caries status was not associated with overall cytokine profiles, but salivary IL-4 was higher in caries-free children.

**Conclusions:** Plasma and salivary cytokines capture largely distinct immune signals in children affected by HIV, and saliva does not appear to serve as a surrogate for plasma with respect to cytokine expression. We did observe that HIV infection and immune status, not dental caries, were the main correlates of cytokine variation in plasma and saliva compartments.

## Introduction

The oral cavity is a key immunologic site where local and systemic immune responses converge, and disruptions in immune homeostasis can manifest as clinically significant disease. In the context of HIV infection, progressive immune dysfunction alters cytokine signaling and may increase susceptibility to opportunistic oral conditions, including oropharyngeal candidiasis, oral hairy leukoplakia,^1,2^ and other inflammatory lesions.^1,3–9^ Dental caries and other common oral diseases are disproportionately prevalent among children living with HIV (HI) and those perinatally-exposed to HIV,^10^ highlighting the broader consequences of immune perturbation during critical developmental periods.

In children living with HIV, HIV infection is most commonly acquired perinatally through mother-to-child transmission, which can occur during pregnancy, childbirth through exposure to maternal blood and vaginal secretions, or postnatally through breastfeeding.^11–14^ HIV primarily targets CD4^+^ helper T cells, using them as sites for replication and proliferation to generate new viral particles. These new virions go on to infect additional CD4^+^ cells, ultimately driving progressive immunosuppression.^15–17^ CD4^+^ T cells are central to adaptive immunity, coordinating immune responses through cytokine secretion and interaction with other immune cells.^18,19^ HIV-mediated depletion of these cells disrupts cytokine production and the regulation of immune-microbiome homeostasis,^20^ which may increase susceptibility to opportunistic infections and contribute to the development of chronic oral diseases like dental caries.

There is a growing body of work that has characterized immune function in the context of both HIV infection and various oral diseases, suggesting that immune profiles may be associated with both oral disease and HIV infection despite ART.^1,2,4,7,8,21–23^ Our team has previously shown that immune status was strongly associated with oral microbial profiles than HIV infection or exposure status.^24^ Together, these studies underscore the importance of elucidating the drivers of immune function and response, and the potential indicators of disease risk.

Saliva is an attractive, non-invasive source of immune biomarkers, and salivary cytokines (soluble proteins secreted by both immune and somatic cells) have been shown to play important roles in immune regulation and the coordination of immune responses. Several studies have measured both plasma and salivary cytokines in the presence of systemic and oral disease. However, the relationship between systemic immune signaling and local, oral immune activity remains poorly defined, especially among children. In particular, it is unclear whether cytokine profiles measured in plasma reflect those observed in saliva, or whether these compartments capture distinct aspects of immune function associated with HIV and its clinical sequelae. Addressing this gap is critical for advancing our understanding of the oral-systemic immune axis and for identifying accessible, non-invasive biomarkers of disease risk.

Multiple studies have explored the use of salivary cytokines as a non-invasive surrogate for systemic immune signaling, which is typically assessed using plasma cytokines.^25–27^ The coordination between local and systemic immune responses has been proposed to result from the diffusion of biomarkers from blood into the oral cavity.^28^ However, this notion is challenged due to the weak plasma-salivary cytokine correlation observed among healthy adults, suggesting that systemic and local immunity are independently regulated in response to separate microenvironments.^25–29^ This underscores the importance of measuring immune biomarkers in both plasma and saliva. To our knowledge, plasma and salivary cytokines have not been compared in matched samples from children perinatally exposed and/or infected with HIV. Assessing both plasma and salivary samples will enable us to observe how systemic and local immune responses may be altered in the context of host clinical phenotypes across children with different HIV exposure and infection status. It may provide immunological insight into the disproportionate oral disease burden among children living with HIV.

In this study, we characterized plasma and salivary cytokine profiles in a cohort of Nigerian children aged 4-11 years, including children living with HIV (HI), HIV-exposed uninfected (HEU), and HIV-unexposed uninfected (HUU) children. Our primary aim was to evaluate the concordance between plasma and salivary cytokine concentrations and characterize differences in cytokine profiles according to HIV infection and exposure status. By evaluating circulating and oral immune mediators within the same participants, this study seeks to clarify the relationship between systemic and oral inflammatory profiles and their potential relevance to dental caries in children affected by HIV.

## Results

### Patient demographics

Matched plasma and saliva samples were analyzed from 467 children: 144 HI, 161 HEU, and 162 HUU children. Sex and mean age were similar across groups (84–87 months). The median CD4^+^ lymphocyte counts differed significantly by HIV status (Kruskal-Wallis test, *p* < 0.001) and was lower in HI children (837 cells/mm³) than in HEU (990 cells/mm³) and HUU children (1,015 cells/mm³; Benjamini-Hochberg [BH]-adjusted p < 0.001). A CD4⁺ T-cell count below 500 cells/mm³ was recorded in 37 HI (25.7%), 6 HEU (3.7%), and 11 HUU (6.8%) children. Out of the 144 children living with HIV, 98 were virally suppressed (<200 copies/mL; 68.0%) and 46 were not suppressed (≥200 copies/mL; 32.0%). Additional characteristics are presented in **Table 1**.

**Table 1:**
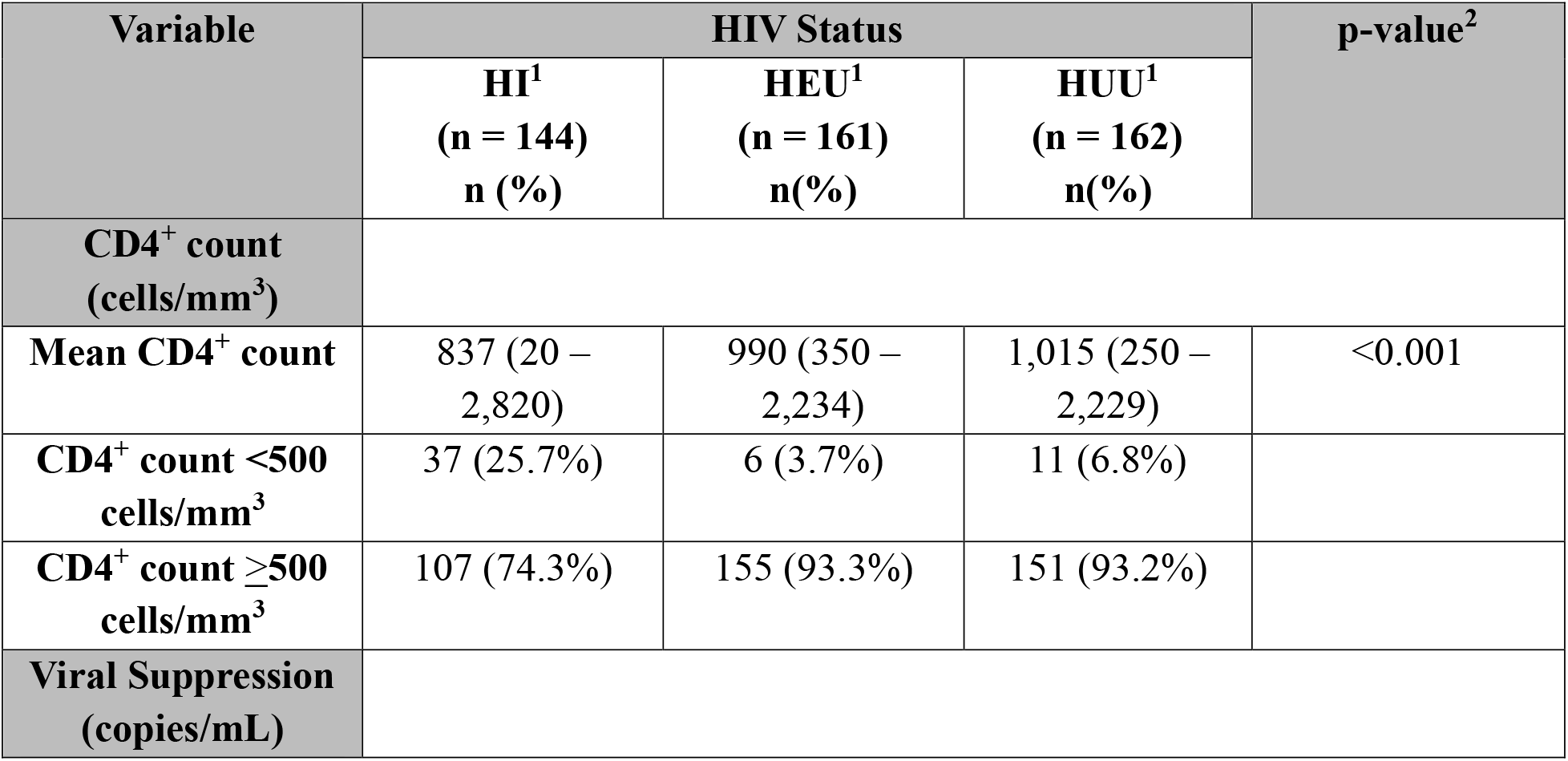
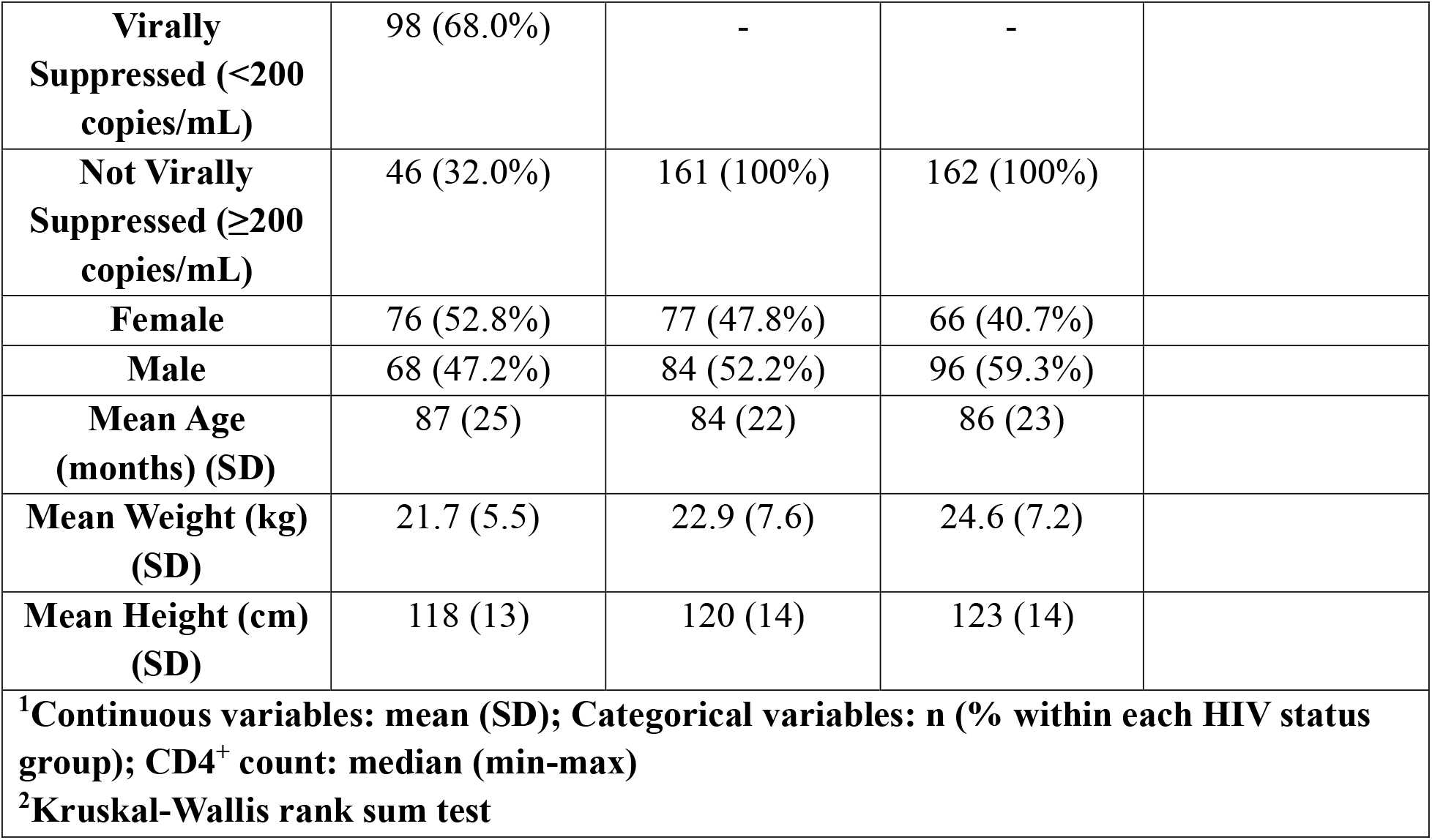
Demographic and clinical characteristics of participants by HIV status. Participants were stratified by HIV status: children living with HIV (HI), HIV-exposed uninfected (HEU), and HIV-unexposed uninfected (HUU). Continuous variables, including age (months), weight (kg), and height (cm), are presented as mean ± standard deviation (SD). Categorical variables, including sex, CD4^+^ T-cell count (≥500 or <500 cells/mm³), and viral suppression status (<200 or ≥200 copies/mL), are presented as the number of participants (n) and percentage (%) within each HIV status group. Median CD4+ T-cell counts, and their ranges, are also provided for each HIV status group.

| **Variable** | **HIV Status** |  |  | **p-value<sup>2</sup>** |
|---|---|---|---|---|
|  | **HI<sup>1</sup><br>(n = 144)<br>n (%)** | **HEU<sup>1</sup><br>(n = 161)<br>n(%)** | **HUU<sup>1</sup><br>(n = 162)<br>n(%)** |  |
| **CD4<sup>+</sup> count (cells/mm<sup>3</sup>)** |  |  |  |  |
| **Mean CD4<sup>+</sup> count** | 837 (20 – 2,820) | 990 (350 – 2,234) | 1,015 (250 – 2,229) | <0.001 |
| **CD4<sup>+</sup> count <500 cells/mm<sup>3</sup>** | 37 (25.7%) | 6 (3.7%) | 11 (6.8%) |  |
| **CD4<sup>+</sup> count ≥500 cells/mm<sup>3</sup>** | 107 (74.3%) | 155 (93.3%) | 151 (93.2%) |  |
| **Viral Suppression (copies/mL)** |  |  |  |  |
| **Virally Suppressed (<200 copies/mL)** | 98 (68.0%) | - | - |  |
| **Not Virally Suppressed (≥200 copies/mL)** | 46 (32.0%) | 161 (100%) | 162 (100%) |  |
| **Female** | 76 (52.8%) | 77 (47.8%) | 66 (40.7%) |  |
| **Male** | 68 (47.2%) | 84 (52.2%) | 96 (59.3%) |  |
| **Mean Age (months) (SD)** | 87 (25) | 84 (22) | 86 (23) |  |
| **Mean Weight (kg) (SD)** | 21.7 (5.5) | 22.9 (7.6) | 24.6 (7.2) |  |
| **Mean Height (cm) (SD)** | 118 (13) | 120 (14) | 123 (14) |  |

**<sup>1</sup>Continuous variables: mean (SD); Categorical variables: n (% within each HIV status group); CD4<sup>+</sup> count: median (min-max)**
**<sup>2</sup>Kruskal-Wallis rank sum test**

### Cytokines are more strongly correlated *within* than *between* plasma and saliva

An initial assessment of cytokine concordance across fluid compartments was performed by calculating pairwise Pearson correlation coefficients (r) between individual cytokines. Correlations among cytokines within the same compartment were also examined to compare within-compartment and between-compartment correlation patterns. Visual inspection of the resulting heatmap suggested that within-compartment correlations were generally stronger, as indicated by larger correlation coefficients, than those observed between plasma and salivary cytokines (**Figure 1**). Correlation coefficients calculated for each HIV-status group gave the same result (**Supplementary Figures 1 A-C**).

**Figure 1:**
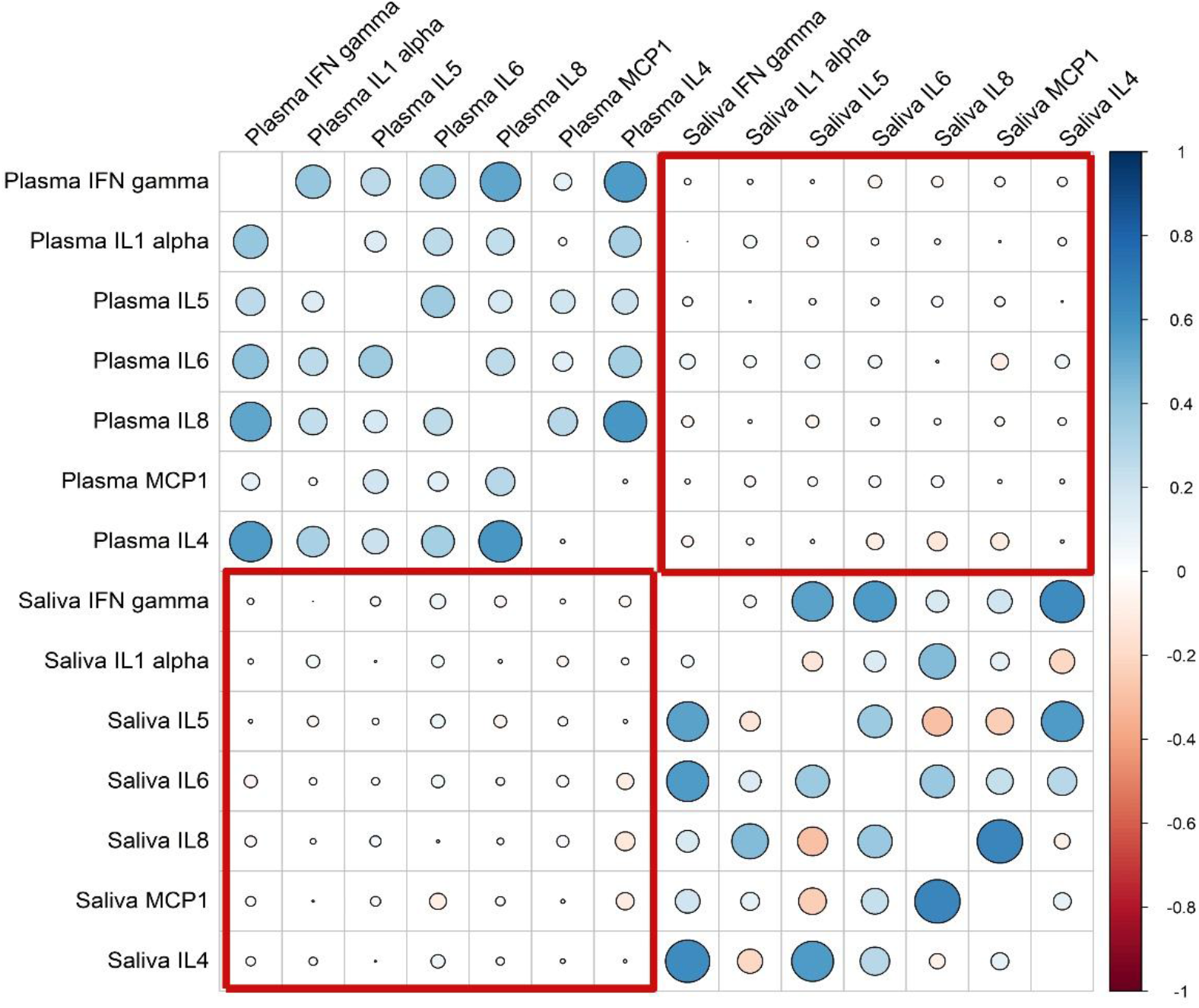
Overall correlations among and between plasma and salivary cytokines. Heatmap illustrating the unadjusted pairwise Pearson correlations (r) among cytokines measured in plasma and saliva. Correlation coefficients are represented by circle size and color, with larger circles indicating stronger correlations. Blue indicates positive correlations, while red indicates negative correlations. The red outlined regions highlight between-compartment cytokine correlations, while the remaining regions represent within-compartment correlations among plasma and salivary cytokines.

### Cytokine profiles are associated with HIV status and CD4⁺ T-cell count but not with caries

To characterize variation in profile-wide systemic and oral immune signaling, multivariate analyses were performed on both plasma and salivary cytokines. Principal component analysis (PCA) was used to visualize patterns of variation in overall plasma and salivary cytokine profiles, while PERMANOVA assessed the associations of HIV status, dental caries status, and CD4^+^ count with cytokine profile composition, adjusting for demographic covariates.

HIV status was significantly associated with overall cytokine profiles in both plasma (p = 0.038; R² = 0.008; **Figure 2A**) and saliva (p < 0.001; R² = 0.017; **Figure 2B**). Pairwise comparisons in plasma suggested differences between HI and HEU children and between HI and HUU children, although neither comparison reached statistical significance (**Table 2A**). In contrast, salivary cytokine profiles differed significantly between HI and both HEU and HUU children (**Table 2B**). CD4^+^ count was significantly associated with plasma cytokine profiles only (p = 0.029; R^2^ = 0.005) and not salivary profiles.

**Figure 2a:**
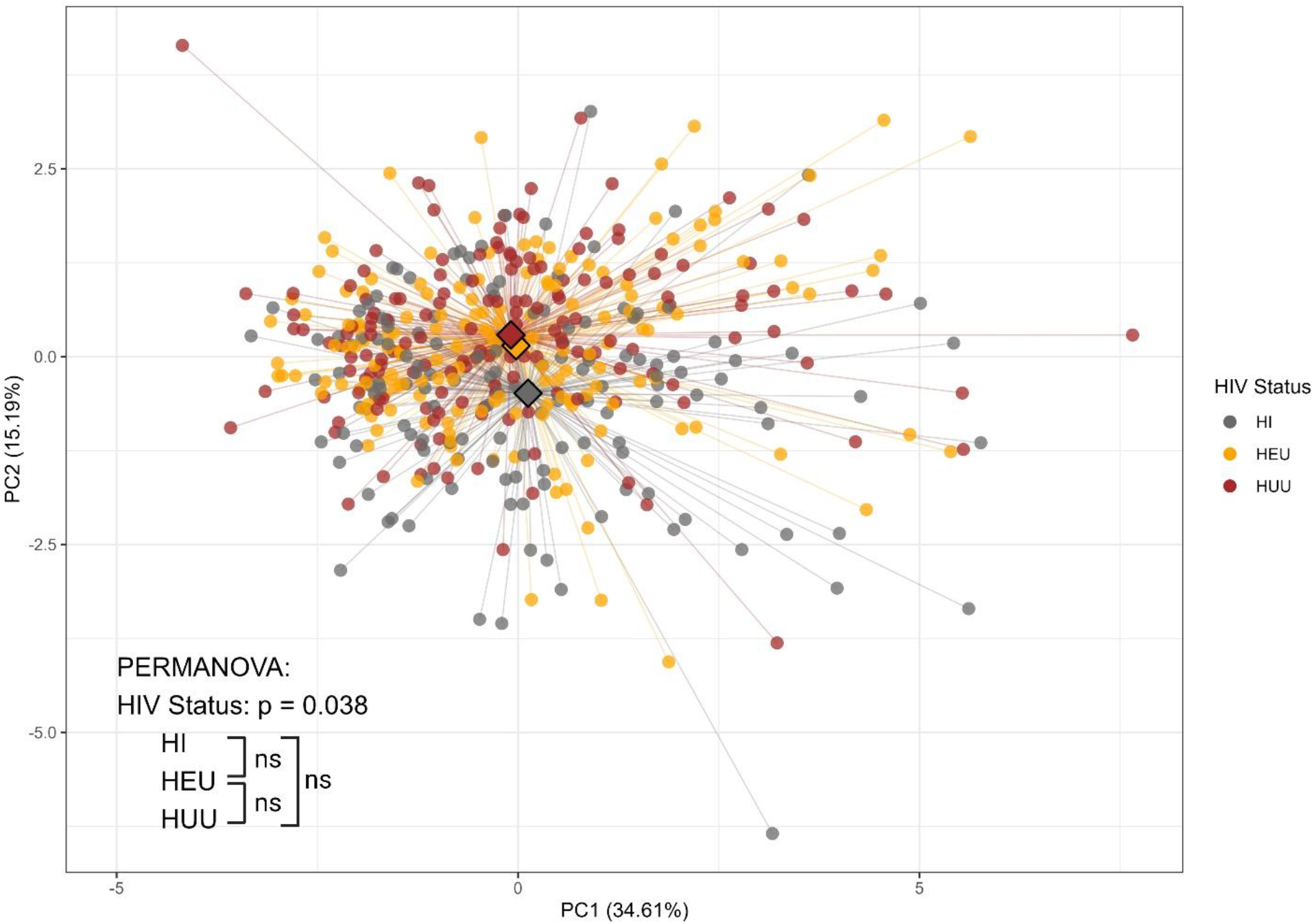
Principal component analysis of plasma cytokine profiles across HIV status groups. PCA plots showing the groupings of cytokine profiles in plasma among children living with HIV (HI), HIV-exposed uninfected (HEU), and HIV-unexposed uninfected children (HUU). Each point represents an individual child plotted in two-dimensional principal component space. Differences in cytokine profiles by HIV status were assessed using PERMANOVA, with nominal p-values reported for each pairwise comparison between groups. “NS” indicates no significant difference; “*” p < 0.05; “**” p < 0.01; and “***” p < 0.001.

**Figure 2b:**
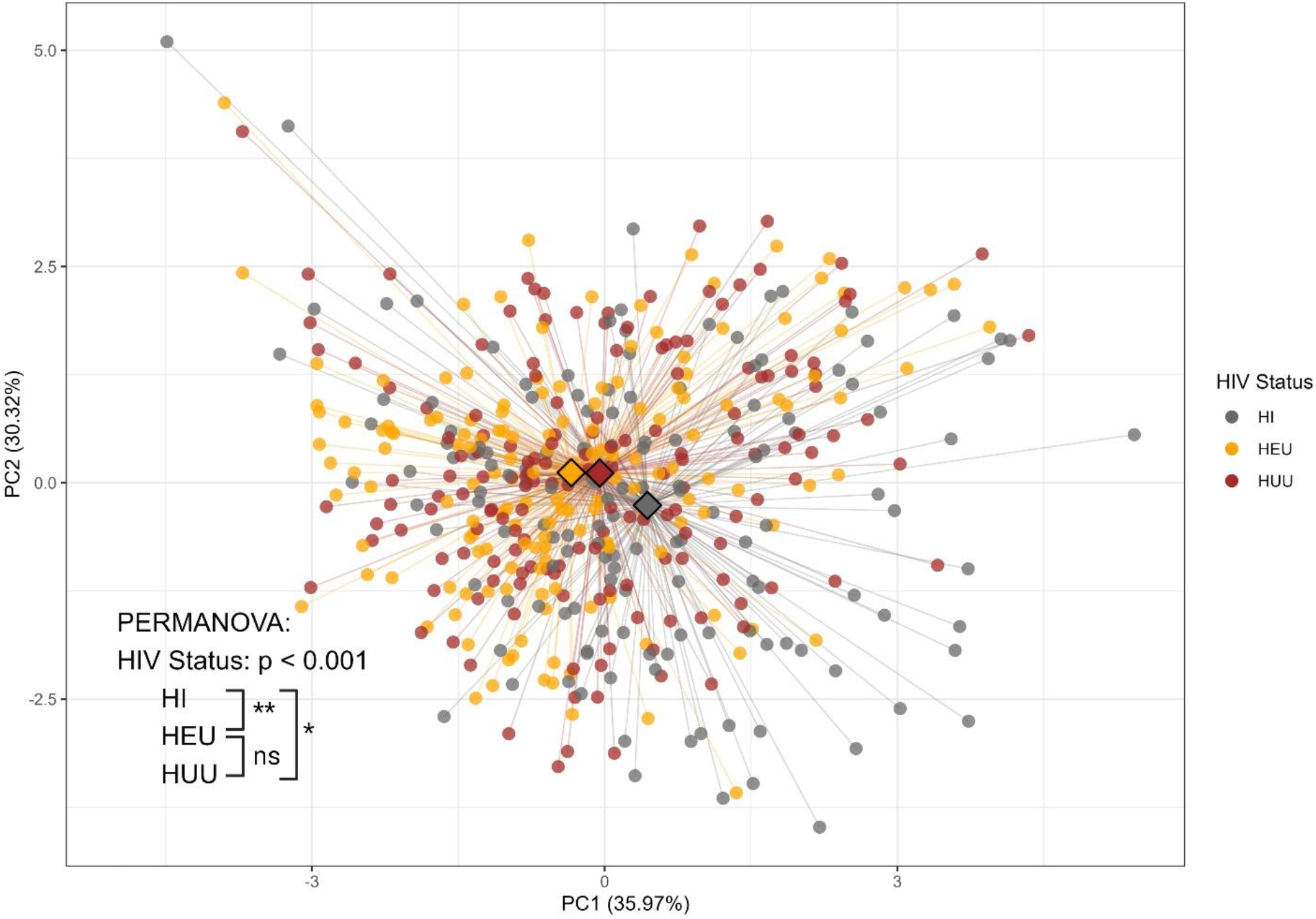
Principal component analysis of salivary cytokine profiles across HIV status groups. PCA plots showing the groupings of cytokine profiles in saliva among children living with HIV (HI), HIV-exposed uninfected (HEU), and HIV-unexposed uninfected children (HUU). Each point represents an individual child plotted in two-dimensional principal component space. Differences in cytokine profiles by HIV status were assessed using PERMANOVA, with nominal p-values reported for each pairwise comparison between groups. “NS” indicates no significant difference; “*” p < 0.05; “**” p < 0.01; and “***” p < 0.001.

**Table 2.**
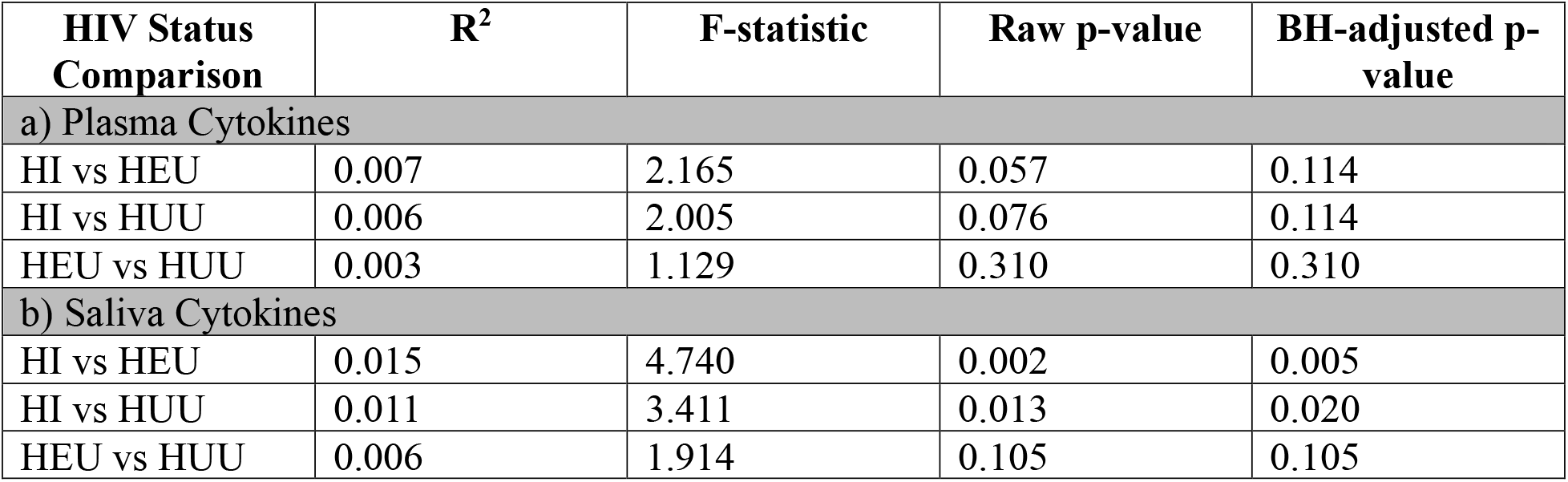
Pairwise PERMANOVA comparisons of plasma and salivary cytokine profiles across HIV status groups. Pairwise differences between HIV status groups in (a) plasma and (b) salivary cytokine profiles, with corresponding PERMANOVA R² values and Benjamini–Hochberg-adjusted p-values for each comparison.

Caries status was not independently associated with overall plasma or salivary cytokine profiles. However, significant associations emerged in plasma when caries status was considered in interaction with HIV status or CD4^+^ count. The HIV x caries interaction approached statistical significance (p = 0.076; R² = 0.007), whereas the CD4^+^ count x caries interaction was significantly associated with plasma cytokine profiles (p = 0.0449; R² = 0.005). Neither interaction was significantly associated with salivary cytokine profiles.

Differences in plasma and salivary cytokine profile dispersion between groups were evaluated using PERMDISP. In plasma, CD4^+^ count groups (≥500 cells/mm^3^ and <500 cells/mm^3^) had different group dispersions (p = 0.009). In saliva, HIV status groups had different group dispersions (p = 0.024). Sensitivity analysis using a subset of children with matching maternal education and employment status did not influence the outcomes of plasma or salivary cytokine profiles. Loadings for principal component 1 (PC1) and principal component 2 (PC2) are presented in **Supplementary Tables 1 and 2**.

### Cytokine Network Organization differs by HIV Status and Biological Compartment

Cytokine correlation network analysis revealed differences in network density across groups, with patterns varying by biological compartment and correlation threshold. Each network comprised seven cytokines, with 21 possible edges. In plasma, HUU children generally exhibited greater network density than HI and HEU children at correlation thresholds of |ρ| ≥ 0.40-0.48, although these differences diminished at more stringent thresholds. In saliva, HEU children exhibited the highest network density at |ρ| ≥ 0.40 (0.381), followed by HI (0.333) and HUU (0.238) (**Table 3A)**. However, salivary network density declined more markedly in HEU children as the threshold increased, whereas HI and HUU networks remained comparatively stable.

**Table 3A:** Cytokine network density across HIV status groups and biological compartments. . Networks comprised seven cytokines (21 possible edges) and were constructed separately for plasma and saliva in HI, HEU and HUU groups. Total edges indicate the number of cytokine pairs meeting the specified absolute correlation threshold (|ρ| ≥ 0.40–0.50), while network density represents the proportion of observed edges among the 21 possible edges. Cytokine associations were adjusted for age and sex. HIV status: children living with HIV (HI), HIV-exposed uninfected (HEU), and HIV-unexposed uninfected (HUU).

| **Plasma** |  |  |  |  |  |  |  |  |  |  |  |  |
| Network parameter | Group | **ρ ≥ 0.40** | **ρ ≥ 0.41** | **ρ ≥ 0.42** | **ρ ≥ 0.43** | **ρ ≥ 0.44** | **ρ ≥ 0.45** | **ρ ≥ 0.46** | **ρ ≥ 0.47** | **ρ ≥ 0.48** | **ρ ≥ 0.49** | **ρ ≥ 0.50** |
|---|---|---|---|---|---|---|---|---|---|---|---|---|
| Total edges | HI | 3 | 3 | 3 | 3 | 3 | 3 | 3 | 3 | 3 | 2 | 1 |
|  | HEU | 3 | 3 | 3 | 3 | 3 | 3 | 3 | 3 | 3 | 3 | 3 |
|  | HUU | 6 | 6 | 6 | 6 | 6 | 6 | 6 | 4 | 4 | 2 | 2 |
| Network density | HI | 0.143 | 0.143 | 0.143 | 0.143 | 0.143 | 0.143 | 0.143 | 0.143 | 0.143 | 0.095 | 0.048 |
|  | HEU | 0.143 | 0.143 | 0.143 | 0.143 | 0.143 | 0.143 | 0.143 | 0.143 | 0.143 | 0.143 | 0.143 |
|  | HUU | 0.286 | 0.286 | 0.286 | 0.286 | 0.286 | 0.286 | 0.286 | 0.19 | 0.19 | 0.095 | 0.095 |
| **Saliva** |  |  |  |  |  |  |  |  |  |  |  |  |
| Network parameter | Group | **ρ ≥ 0.40** | **ρ ≥ 0.41** | **ρ ≥ 0.42** | **ρ ≥ 0.43** | **ρ ≥ 0.44** | **ρ ≥ 0.45** | **ρ ≥ 0.46** | **ρ ≥ 0.47** | **ρ ≥ 0.48** | **ρ ≥ 0.49** | **ρ ≥ 0.50** |
| Total edges | HI | 7 | 6 | 6 | 6 | 6 | 6 | 6 | 5 | 5 | 5 | 5 |
|  | HEU | 8 | 7 | 7 | 7 | 6 | 5 | 5 | 3 | 3 | 2 | 2 |
|  | HUU | 5 | 5 | 5 | 5 | 5 | 5 | 5 | 5 | 5 | 5 | 5 |
| Network density | HI | 0.333 | 0.286 | 0.286 | 0.286 | 0.286 | 0.286 | 0.286 | 0.238 | 0.238 | 0.238 | 0.238 |
|  | HEU | 0.381 | 0.333 | 0.333 | 0.333 | 0.286 | 0.238 | 0.238 | 0.143 | 0.143 | 0.095 | 0.095 |
|  | HUU | 0.238 | 0.238 | 0.238 | 0.238 | 0.238 | 0.238 | 0.238 | 0.238 | 0.238 | 0.238 | 0.238 |

In contrast to the within-compartment cytokine networks observed in plasma and saliva, no inter-compartmental (plasma-saliva) connections were identified across the three HIV status groups when applying the same correlation magnitude thresholds (|ρ| ≥ 0.40-0.48). The maximum absolute correlation coefficients for plasma-saliva cytokine pairs were relatively weak in all groups: HI (0.16), HEU (0.17), and HUU (0.22) **(Table 3B)**. These findings suggest limited correlation-based connectivity between circulating and salivary cytokine profiles, despite the presence of stronger associations within each biological compartment.

**Table 3B.** Cross-compartmental plasma–saliva cytokine correlations across HIV status groups. . Partial Spearman correlations were calculated for 49 plasma–saliva cytokine pairs (7 × 7). Maximum absolute correlation coefficients (|ρ|), P values and FDR-adjusted P values are reported. No associations remained significant after FDR correction. HIV status: children living with HIV (HI), HIV-exposed uninfected (HEU), and HIV-unexposed uninfected (HUU).

| **Group** | **Maximum \|ρ\|** | **P value** | **FDR** |
|---|---|---|---|
| HI | 0.16 | 0.058 | 0.901 |
| HEU | 0.17 | 0.031 | 0.97 |
| HUU | 0.22 | 0.004 | 0.23 |

Node-level centrality analysis revealed compartment-specific patterns across HIV status groups (**Supplementary Table 3**). In plasma, IFN-γ was prominent in HI and HUU, exhibiting the highest eigenvector centrality (1.00 in both groups) and strength (1.59 and 2.40, respectively), whereas IL-4 was most central in HEU, with the highest strength (2.07), betweenness (0.40), closeness (0.31) and eigenvector centrality (1.00). In saliva, IL-6 and IL-8 were prominent in HI, with IL-6 exhibiting the highest betweenness (0.33) and closeness (0.36), and IL-8 exhibiting the highest strength (2.28). In HEU and HUU saliva networks, IL-5 was the most central cytokine, exhibiting the highest degree (6.00 in both groups), strength (2.41 and 2.37, respectively), betweenness (0.47 and 0.33, respectively) and eigenvector centrality (1.00 in both groups).

### HIV Status and CD4^+^ Count are Associated with Variation in Plasma and Salivary Cytokines

Separate multivariable linear regression models assessed associations of individual plasma and salivary cytokines with HIV status, CD4^+^ count, caries status, current weight (kg), height (cm), age (months), sex, and breast-feeding duration (months). HIV status and CD4^+^ count were the predominant variables significantly associated with cytokine concentrations in both compartments.

HIV status was significantly associated with 4 plasma and 3 salivary cytokines. In plasma, the HI group had higher IL-5, MCP-1, and TNF-α concentrations than HUU children (**Table 4**). Compared with HEU children, the HI group had higher concentrations of IL-5, IL-6, and MCP-1. In saliva, HEU children had lower IL-6 concentrations than HUU children. The HI group had higher IL-6 and MCP-1 concentrations than HUU children, and higher IFN-γ, IL-6, and MCP-1 concentrations than HEU children (**Table 5**). All comparisons remained significant after BH adjustment.

**Table 4.** Multivariable linear regression of plasma cytokine concentrations. Significant associations of HIV status, CD4^+^ count, age (months), weight (kg), height (cm), sex, and breastfeeding duration (months) with plasma cytokine concentrations. R^2^ indicates the proportion of variability in cytokine concentrations explained by the predictors in each regression model. HIV-status comparisons include HI vs HUU, HEU vs HUU, and HI vs HEU. Benjamini–Hochberg (BH) correction was applied to HIV-status comparisons; other predictors are reported using unadjusted p-values. Only significant associations are shown. HIV status groups: children living with HIV (HI); HIV-exposed uninfected (HEU); HIV-unexposed uninfected (HUU).

| **Cytokine** | **Predictor/Comparison** | **Estimate** | **Std. error** | **p-value** | **BH-adjusted p-value** | **Partial R<sup>2</sup>** |
|---|---|---|---|---|---|---|
| IL-5 | **HIV Status**: |  |  |  |  |  |
|  | HI vs HUU | 0.393 | 0.131 | 0.00288 | 0.00865 | 0.024 |
|  | HI vs HEU | 0.361 | 0.134 | 0.00721 | 0.0108 | 0.024 |
| IL-6 | **HIV Status**: |  |  |  |  |  |
|  | HI vs HEU | 0.444 | 0.159 | 0.00557 | 0.0167 | 0.019 |
|  | **CD4<sup>+</sup> count (cells/mm<sup>3</sup>)**: |  |  |  |  |  |
|  | Per 1-cell/mm<sup>3</sup> increase | -4.7 x 10<sup>-4</sup> | 0.000 | <0.001 | - | 0.025 |
|  | **Current Height (cm)**: |  |  |  |  |  |
|  | Per 1-cm increase | 0.020 | 0.009 | 0.0241 | - | 0.011 |
| MCP-1 | **HIV Status**: |  |  |  |  |  |
|  | HI vs HUU | 0.346 | 0.092 | <0.001 | <0.001 | 0.034 |
|  | HI vs HEU | 0.269 | 0.094 | 0.00421 | 0.00631 | 0.034 |
|  | **CD4<sup>+</sup> count (cells/mm<sup>3</sup>)**: |  |  |  |  |  |
|  | Per 1-cell/mm<sup>3</sup> increase | -2.67 x 10<sup>-4</sup> | 0.000 | <0.001 | - | 0.024 |
| TNF-α | **HIV Status**: |  |  |  |  |  |
|  | HI vs HUU | 0.432 | 0.173 | 0.013 | 0.0389 | 0.015 |
| IL-1β | **Current Height (cm)**: |  |  |  |  |  |
|  | Per 1-cm increase | 0.026 | 0.012 | 0.0325 | - | 0.010 |
| IL-4 | **Sex**: |  |  |  |  |  |
|  | Female vs Male | -0.239 | 0.100 | 0.0172 | - | 0.012 |

**Table 5.**
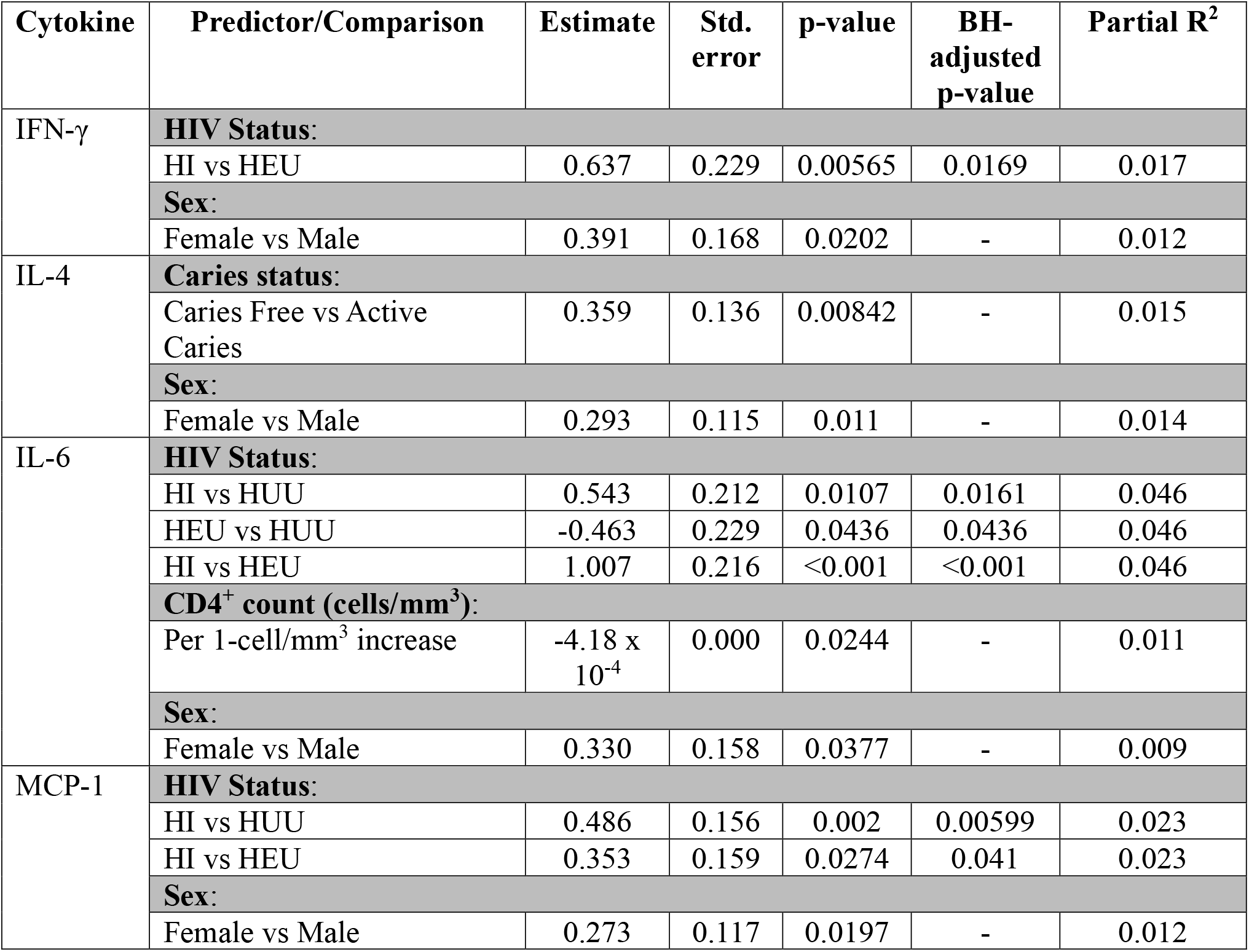
Multivariable linear regression of salivary cytokine concentrations. Significant associations of HIV status, CD4^+^ count, age (months), weight (kg), height (cm), sex, and breastfeeding duration (months) with salivary cytokine concentrations. R^2^ indicates the proportion of variability in cytokine concentrations explained by the predictors in each regression model. HIV-status comparisons include HI vs HUU, HEU vs HUU, and HI vs HEU. Benjamini–Hochberg (BH) correction was applied to HIV-status comparisons; other predictors are reported using unadjusted p-values. Only significant associations are shown. HIV status groups: children living with HIV (HI); HIV-exposed uninfected (HEU); HIV-unexposed uninfected (HUU)

Higher CD4^+^ count was significantly associated with lower plasma IL-6 (β = −4.70 × 10⁻⁴ per cell/mm³) and plasma MCP-1 (β = −2.67 × 10⁻⁴ per cell/mm³) (**Table 4**), and with lower salivary IL-6 (β = −4.18 × 10⁻⁴ per cell/mm³) (**Table 5**).

Caries was significantly associated with only one salivary cytokine; salivary IL-4 was higher in caries-free children than in children with active caries (β = 0.359, p = 0.008) (**Table 5**). Current height and sex were also associated with plasma and salivary cytokine concentrations in the cohort; however, neither consistently influenced the same cytokine in both fluid compartments (except between sex and IL-4, but their directions were opposites; **Table 4-5**). Sensitivity analysis using a subset of children with matching maternal education and employment status did not influence the outcomes of plasma or salivary cytokine multivariable linear models.

In a subset of 203 children with available salivary flow rate data, multivariable linear regression revealed significant inverse associations between salivary flow rate and salivary concentrations of IL-8 (p = 0.006) and MCP-1 (p < 0.001) (**Supplementary Table 4**).

### Caries-cytokine associations differ by HIV status and CD4+ T-Cell count

Caries interaction models with HIV and CD4^+^ count were fitted to investigate if there are significant interactions between caries and either HIV status or CD4^+^ count. Significant interactions between HIV exposure/infection status and caries status were observed for plasma IL-1α, with both HI and HUU differing significantly from HEU (**Supplementary Table 5a**). Evidence of interaction was more limited for plasma IL-1β, with a significant interaction observed only between HI and HEU groups in relation to caries status (**Supplementary Table 5a**). Stratified estimated marginal means comparisons showed that active caries was associated with lower IL-1α concentrations among HEU children (β = −1.253; BH-adjusted p = 0.008) (**Supplementary Table 5b**). In contrast, higher concentrations of IL-1α were observed among HI and HUU children with active caries, although these differences were not statistically significant (BH-adjusted p > 0.05) (**Supplementary Table 5b**). A similar trend was observed for IL-1β, but none of the caries comparisons reached statistical significance (BH-adjusted p > 0.05) (**Supplementary Table 5b**). For both IL-1α and IL-1β, only a few direct comparisons of caries-associated differences demonstrated some differences between HI and HEU, but they failed to reach statistical significance in either caries status (**Supplementary Table 5c**).

A significant interaction was observed between CD4^+^ count and caries status in the plasma IL-1α (β = 0.001; p = 0.011) and IFN-γ (β = 7.567 x 10^-4^; p = 0.029) models (**Supplementary Table 5d**). Stratified analyses showed a significant inverse association between CD4^+^ count and IL-1α concentrations among children with active caries (β = −1.33 x 10^-3^; 95% CI: −2.30 x 10^-3^ –-3.70 x 10^-3^, p = 0.007). Among caries-free children, the positive association between CD4^+^ count and IFN-γ was weak and borderline significant (β = 3.00 x 10^-4^, 95% CI: −2.44 x 10^-5^ – 6.24 x 10^-4^, BH-adjusted p = 0.070) (**Supplementary Table 5e**).

## Discussion

To our knowledge, this is among the first studies to characterize both plasma and salivary cytokine profiles in matched samples from children with perinatal HIV infection or exposure. There were three main findings from this investigation. First, cytokines were more strongly correlated within each fluid compartment but not between fluids. Second, HIV infection was associated with cytokine profiles in both fluids, more clearly in saliva, and CD4⁺ T-cell count was associated with plasma profiles and with IL-6 in both fluids. Third, caries status was not associated with overall cytokine profiles; its associations were limited to salivary IL-4 and to plasma IL-1α, IL-1β, and IFN-γ in interaction with immune phenotypes.

Saliva has been proposed as a non-invasive surrogate for plasma on the premise that circulating cytokines can translocate to the oral cavity.^27,28^ For this cohort of children, our data does not support this. Within-fluid correlations were moderate, whereas the strongest plasma-saliva correlation in any HIV-status group was 0.22. This aligns with several studies in adults, in which plasma-saliva correlations were absent or were modest and confined to a few cytokines^25,28,29^, and with the weak relationship reported between salivary cytokines and plasma measures in adults with HIV.^22^ Salivary cytokines are produced locally by salivary glands, oral epithelium, and resident and recruited immune cells, and they respond to local conditions such as oral hygiene, gingival inflammation, plaque accumulation, dental caries, and the oral microbiota.^29,30^ The compartmentalization observed in our study was evident across all three HIV-status groups. Consistent with this finding, node-level centrality analysis revealed distinct patterns of cytokine network organization between plasma and saliva. IFN-γ and IL-4 were prominent in plasma networks, whereas IL-5, IL-6 and IL-8 occupied prominent positions in salivary networks, with these patterns also varying by HIV status. Network density also varied by HIV status within each compartment, although these differences were not consistent across correlation thresholds, highlighting the need for further validation. Together, these findings indicate that plasma and salivary cytokines should be measured concurrently rather than treated as interchangeable indicators of inflammatory status.

Previous studies using multivariate methods have shown that plasma cytokine and chemokine profiles differ between people with HIV and uninfected controls irrespective of virologic status.^31,32^ For this study, we wanted to extend this approach to saliva and to HEU children, a unique but growing population. HIV status was associated with overall profiles in both fluids, but it explained little of the variation (R² < 0.02), and pairwise differences between HI children and the two uninfected groups were significant only in saliva. Because dispersion also differed between HIV-status groups in saliva, part of this difference may reflect greater heterogeneity among HI children rather than a uniform shift. The salivary difference may reflect both the systemic effects of HIV on immune activation and HIV-associated changes in the oral environment, including altered salivary properties.^33^ CD4⁺ T-cell count was associated with plasma but not salivary profiles, consistent with a closer relationship between systemic immune status and circulating cytokines.

After examining differences in multivariate systemic and local cytokine profiles across HIV status groups, we used multiple linear regression models to assess the associations of individual cytokines with HIV groups while adjusting for breast-feeding duration, age, weight, height, and sex. We observed that some differences were specific to one fluid compartment. Plasma IL-5 was higher in HI than in HEU and HUU children, plasma TNF-α was higher in HI than in HUU children, and salivary IFN-γ was higher in HI than in HEU children. Notably, IL-6 and MCP-1 behaved similarly in both fluids. IL-6 was higher in HI than in HEU children in plasma and saliva; in saliva, it was also higher in HI than in HUU children and lower in HEU than in HUU children. MCP-1 was higher in HI children than in both uninfected groups in both fluids. Higher CD4⁺ T-cell count was associated with lower IL-6 in both fluids and with lower MCP-1 in plasma only. These parallel group differences occurred even though concentrations of the same cytokine were not correlated between fluids within individuals, which suggests that HIV infection might act on both compartments through separate local and systemic sources rather than through transfer from blood to saliva.

The lower salivary concentration of IL-6 in HEU compared to HUU children was unexpected. Reports of children in the HEU group describe both attenuated and enhanced cytokine responses depending on the stimulus and age,^34,35^ and recent cohorts found lower concentrations of several circulating cytokines in HEU compared to HUU that persisted beyond infancy.^36,37^ Our finding is in the same direction but was limited to one cytokine in one fluid and needs replication. Also, given we enrolled from a hospital setting, it is possible that HUU children might not reflect a truly healthy unexposed group compared to HEU. This is plausible because in this cohort, we have also observed a lower though non-significant odds of having carious lesions compared to HUU children.

Caries status had little independent association with cytokines. Salivary IL-4 was higher in caries-free children, but this was a single association while adjusting for breast-feeding duration, age, weight, height, and sex. In plasma, the association between caries and IL-1α depended on HIV status. We observed that caries was associated with lower IL-1α in HEU children, whereas IL-1α tended to be higher with caries burden in HI and HUU children, although neither within-group difference was significant. IL-1β showed a weaker pattern in the same direction.

In our cohort, salivary IL-6 varied with HIV status and CD4⁺ T-cell count. The importance of IL-6 is reflected in multiple studies, where IL-6 was found to be a prevalent salivary cytokine among children and young adults with dental caries.^38^ Some have suggested that an increase in salivary IL-6 concentration is an indicator of the presence and severity of dental caries, serving key roles in the body’s response to infections by cariogenic bacteria (i.e. *Streptococcus mutans* and *Lactobacillus*).^38–40^ For example, IL-6 is found to be produced by activated macrophages, monocytes, and fibroblasts in inflamed tissues.^40^ In addition, IL-6 was increased among children with early childhood caries (ECC) compared to those without any caries in saliva; however, such observations were made among children without any systemic, chronic diseases.^41,42^ Unlike these previous studies, we did not observe significant associations between IL-6 and dental caries in either plasma or saliva. This may be partly explained by the stronger association of HIV status with cytokine levels in our cohort, potentially overshadowing associations with caries. Additionally, these previous studies excluded children with systemic/chronic illnesses. A future goal would be to investigate whether the upregulation in plasma and salivary IL-6 increases the likelihood of developing dental caries in children living with HIV. In terms of salivary MCP-1 levels, it was previously observed to be elevated among individuals with severe chronic periodontitis compared to healthy controls. MCP-1 has been implicated in the chemoattraction of monocytes to sites of inflammation.^43^

Together, these findings indicate that cytokine variability across HIV status and CD4^+^ count may not entirely be uniform, and may differ by cytokine, biological compartment, and other demographic and clinical factors. This complexity is consistent with previous studies associating salivary cytokine expression with HIV status and CD4^+^ counts in the presence of opportunistic infections.^7,21,44^ Viral load may also influence salivary cytokine expression among people living with HIV,^1,2^ while antiretroviral therapy (ART), periodontitis, and CD4^+^ count have been reported to contribute to variations in salivary cytokine profiles.^1,5,7,21–23,44^ These findings suggest that HIV-associated cytokine profiles vary according to clinical presentation and other contributing factors, even among individuals receiving ART. Salivary rate is another important factor, as higher flow rates have been associated with lower concentrations of IL-1α, IL-1β, IL-6, IL-8, IL-10, and TNF-α, potentially reflecting a dilution effect among salivary cytokines.^30^ However, in our main analyses, we did not normalize cytokine concentrations based on salivary flow rate because flow rates differ between HUU and HEU children in our cohort.^30^ As HIV status may influence both salivary flow rate and cytokine concentrations, normalization could attenuate biologically relevant differences between groups. Therefore, cytokine concentrations were analyzed without flow-rate adjustment, and the findings should be interpreted in the context of the clinical and physiological factors that may contribute to variation across biological compartments.

Although the proposed biological mechanisms for caries development can vary, a common theme here is that alterations in cytokine expression may create conditions that favor the manifestation of opportunistic infections and oral pathology. By reducing CD4^+^ T-cell numbers and altering cytokine production, HIV may shift this balance and alter immune function within the oral cavity.^1,2,4,7,21,44,45^ Consistent with this hypothesis, the present study does demonstrate that both HIV status and CD4^+^ count can influence both plasma and salivary cytokine expression.

Despite a higher prevalence of dental caries among children living with HIV, caries status was not significantly associated with overall multivariate cytokine profiles, neither was it widely associated with individual cytokines in plasma or saliva. However, associations between caries and individual plasma cytokines varied according to HIV exposure/infection status and CD4^+^ T-cell count. Our group previously reported associations between dental caries and inflammatory biomarkers in the context of perinatal HIV exposure,^46^ the present findings suggest that these relationships may be influenced by the underlying immunological context.

Several limitations warrant consideration. First, the selected cytokine panel does not capture the full complexity of immune signaling. Measurements calculated from this panel on whole saliva and plasma samples reflect aggregate cytokine activity rather than responses from specific cell populations. Although the manufacturer indicates that the analytes are detectable in saliva, independent validation of assay performance in this biological matrix was not performed. Multiplex bead-based assays may also have lower analytical sensitivity than single-analyte immunoassays, particularly at low cytokine concentrations. Second, potential assay-related batch effects cannot be excluded, as between-run variability was not directly assessed or accounted for across all 467 participants. Although intra-assay CVs remained below 15% in both biological matrices, greater inter-assay variability suggests that differences between assay runs may have influenced cytokine measurements. Nevertheless, adjusting for batch effects using ComBat reduced inter-assay variability, allowing our primary results to be relatively robust. Accordingly, some of the profile patterns observed may reflect technical variation in addition to underlying biological differences.

In conclusion, plasma and salivary cytokines captured largely distinct immune signals in this cohort of children aged 4–11 years, and saliva does not appear to be a surrogate for plasma cytokine measurement in children affected by HIV. HIV infection and CD4^+^ T-cell count were the main correlates of cytokine variation, although they explained only a small proportion of the variation in overall profiles. IL-6 and MCP-1 were the cytokines most consistently elevated in children living with HIV across both compartments, and overall cytokine profiles distinguished these children from HEU and HUU children more clearly in saliva than in plasma. Caries status was not associated with overall cytokine profiles. However, its associations with IL-1α, IL-1β, and IFN-γ varied by HIV exposure/infection status and CD4+ T-cell count, but these exploratory findings require confirmation. Together, our findings underscore the importance of assessing systemic and oral immune compartments in parallel when studying oral disease risk in children affected by HIV. Future longitudinal studies are needed to determine whether these cytokine alterations causally lead to dental caries development in HI and HEU children rather than reflecting differences in HIV exposure, immune status, and oral health.

## Materials and Methods

### Study Design

This cross-sectional analysis used samples and data from children (N=467; aged 4-11 years) from the Dental Caries and its Association with the Oral Microbiomes and HIV in Young Children-Nigeria (DOMHaIN) cohort^47^ at the University of Benin Teaching Hospital (UBTH) between May and November 2019. Participants were recruited at UBTH by trained healthcare staff and were categorized into three groups: children living with HIV (HI), HIV-exposed but uninfected (HEU), and HIV-unexposed and uninfected (HUU). HI and HEU participants were recruited from the Pediatric Special Treatment HIV Clinic, while HUU children were recruited from the Pediatric Out-Patient Clinic. Groups comprised of 144 HI, 161 HEU, and 162 HUU children. Sociodemographic, feeding, and oral hygiene data were obtained with standardized caregiver questionnaires.

The guidelines governing the administration of ART among children living with HIV participating in this study follow the protocols outlined by the Nigerian Federal Ministry of Health. ART was initiated for all children living with HIV at the time of recruitment from UBTH and have remained on ART through the duration of the study. ART regimens provided to these children consisted of a combination of different inhibitor drug classes. Children in the HI group were given a Nucleoside/Nucleotide reverse Transcriptase Inhibitor (NRTIs) in combination with a Non-Nucleoside Reverse Transcriptase Inhibitor (NNRTIs), Protease Inhibitor (PI), and/or Integrase Strand Transfer Inhibitor (INSTI) according to the recommended ascribed first-line regimens outlined for children <3 yrs and children 3 - <10 yrs (**Table 6**).^48^

**Table 6:** Antiretroviral therapies administered to children living with HIV (HI). A frequency table listing the number of children living with HIV receiving each ART treatment as recommended by the Nigerian Federal Ministry of Health. The abbreviations are shorthand for the name of the drug administered. Nucleoside Reverse Transcriptase Inhibitors (NRTIs): Abacavir (ABC), Tenofovir Disoproxil Fumarate (TDF), Lamivudine (3TC), Emtricitabine (FTC), Zidovudine (AZT). Non-Nucleoside Reverse Transcriptase Inhibitor (NNRTIs): Efavirenz (EFV) and Nevirapine (NVP). Integrase Strand Transfer Inhibitor (INSTI): Dolutegravir (DTG). Protease Inhibitor (PI): Lopinavir/Ritonavir (LPV/r). Only 2 children were omitted from this table due to missing ART therapy information.

| **ART Treatment** | **Number of Children** | **Percentage (%)** |
|---|---|---|
| AZT/3TC/NVP | 83 | 58.45 |
| AZT/3TC/EFV(NVP) | 26 | 18.31 |
| ABC/3TC/LPV/r | 8 | 5.63 |
| TDF/3TC/DTG | 7 | 4.93 |
| AZT/3TC/EFV | 6 | 4.23 |
| ABC/3TC/DTG | 5 | 3.52 |
| ABC/3TC/NVP | 2 | 1.41 |
| ABC(TDF)/3TC/LPV/r | 1 | 0.70 |
| ABC/3TC/EFV | 1 | 0.70 |
| ABC/FTC/EFV | 1 | 0.70 |
| AZT/3TC/EFV/NVP | 1 | 0.70 |
| TDF/3TC/LPV/r | 1 | 0.70 |

### Sample Collection

All efforts were made to ensure that biospecimen collection and handling procedures were conducted aseptically. Blood and saliva samples are collected to measure the concentration of dissolved cytokines found in plasma and saliva (oral cavity) respectively.

#### Blood/Plasma

At enrollment, 10 mL of venous blood was collected from each child by venipuncture. Plasma was separated by centrifugation at 4000 rpm revolution for 10 mins, and the clear plasma separated and stored for future cytokine profile assays within 1 hour of collection, aliquoted, and stored at −80 °C. Plasma aliquots were stored and subsequently shipped on dry ice to the US for further processing as described below.

#### Saliva

Unstimulated whole saliva was collected at the same visit as the blood sample. The child laid on a dental chair with the head raised, saliva was allowed to pool in the mouth for 5 minutes and was then aspirated with sterile plastic Pasteur pipettes into two pre-labelled 2 mL cryogenic vials kept on ice. Samples were stored at −80 °C and shipped as described for plasma. Processing of these samples are previously outlined in the study protocol.^47^ Salivary flow rate was measured by dividing the recorded volume of saliva collected by the duration of saliva production, typically five minutes. These calculated values were then documented as the salivary flow rate for each participant in the study.

Before the assays, thawed plasma and saliva samples were brought to room temperature and centrifuged at 2,000 × *g* for 5 minutes. Plasma supernatants were used directly for the assay. Saliva supernatants were treated with protease inhibitor cocktail at a 1:500 volume ratio (protease inhibitor cocktail to saliva), centrifuged at 10,000 rpm for 10 minutes, and diluted 1:2 with the kit-provided assay buffer.

### Milliplex Multiplex Immunoassays

Plasma and salivary cytokine concentrations were quantified using the MILLIPLEX Human Cytokine/Chemokine/Growth Factor Panel A Magnetic Bead Assay (MilliporeSigma), which measures 11 cytokines: IFN-γ, IL-1α, IL-1β, IL-2, IL-4, IL-5, IL-6, IL-8, IL-10, MCP-1, and TNF-α. The assay was performed according to the manufacturer’s protocol, with samples analyzed in duplicate and the mean concentration calculated.

#### Assay precision

Mean intra-assay coefficients of variation (CVs) ranged from 6.13% to 11.08% in plasma and 5.56% to 10.44% in saliva, below the recommended 15% threshold (**Supplementary Table 6-7**). ComBat (*BatchCorrector*) was used to help minimize/eliminate inter-assay related batch effects.^49^ Cytokine concentrations for the standards were log_2_(x+1) transformed prior to ComBat adjustment, then back transformed after adjustment.^49^ Inter-assay precision was more variable (**Supplementary Table 8**). In plasma and saliva, IL-1α, IL-10, MCP-1, and TNF-α are the only cytokines with batch-adjusted inter-assay CV% >20%. All other cytokines had a batch-adjusted inter-assay CV% <20%.

#### Data processing

Following quantification using the Luminex xMAP Intelliflex system, cytokine concentrations below the lower limit of detection (LLOD) were imputed as ½ the cytokine-specific LLOD, defined as the minimum detectable concentration (MinDC) + 2 standard deviations (SD) for the overnight incubation protocol.^50^ Plasma IL-2 and salivary IL-1β, IL-2, IL-10, and TNF-α were excluded from the analyses because more than 30% of their measured concentrations fell below the LLOD (**Supplementary Table 9**). This 30% threshold was used in other studies.^29,30^ Cytokine concentrations were then adjusted for batch effects using ComBat. Raw cytokine concentrations were log_2_(x+1) transformed prior to ComBat adjustment, then back transformed after adjustment.^49^ The adjusted cytokine distributions were assessed using boxplots. Because adjusted concentrations were highly right-skewed, values were log_2_(x+1)-transformed to reduce right skewness and accommodate zero or near-zero values.^49,51,52^

### HIV status and viral load

HIV infection and exposure status were determined from the medical records of the mother and child and confirmed by HIV testing of the child at enrollment.^47^ Serial rapid antibody tests were used: Determine as the first test, Uni-Gold to confirm a reactive result, and Stat-Pak as the tie-breaker for discordant results.^53^ For HEU children, records of early infant diagnosis by HIV DNA PCR (COBAS AmpliPrep/COBAS TaqMan HIV-1 Qualitative Test, version 2.0) were reviewed; under national guidelines this test is performed at 6 weeks of age, at 4 months for non-breastfed infants, or 2 months after cessation of breastfeeding.^48,54^ HIV viral detection and viral load quantification were performed by UBTH laboratory technicians using Real-Time Polymerase Chain Reaction (RT-PCR). Viral suppression was determined based on a cutoff of 200 copies/mL. Viral suppression is any viral load <200 copies/mL, while ≥200 copies/mL is considered not virally suppressed. This cutoff had been used in previous studies.^55,56^

Antiretroviral therapy (ART) followed the guidelines of the Nigerian Federal Ministry of Health.^48^ All HI children were receiving ART at enrollment and remained on ART during the study. Regimens combined a nucleoside or nucleotide reverse transcriptase inhibitor backbone with a non-nucleoside reverse transcriptase inhibitor, a protease inhibitor, or an integrase strand transfer inhibitor (**Table 6**).

### Quantification of CD4 and CD8 lymphocytes

CD4⁺ and CD8⁺ T-cell counts were measured for all children (regardless of group) by flow cytometry on a Sysmex/Partec CyFlow Counter II with fluorescently labelled monoclonal antibodies and a 532 nm solid-state laser.^57,58^ CD4⁺ T-cell count was analyzed as a continuous variable (cells/mm³) and, for dispersion analyses, in two groups (≥500 and <500 cells/mm³).

### Oral health examination

The presence and absence of dental caries were evaluated during a dental examination performed by a qualified dentist who was unaware of the HIV exposure status of the child whom they are examining. The WHO criteria for the assessment of caries and International Caries Detection and Assessment (ICDAS) scores were used to determine if the child’s dentition had active caries (CA) or were caries-free (CF).^59,60^ Children were classified as having active caries or as caries-free.

### Statistical Analysis

All analyses were performed in R (version 4.2.1). Multiple linear regression assessed associations of HIV status, CD4⁺ T-cell count (cells/mm³), and caries status in the presence of other covariates (sex, age (months), height (cm), weight (kg), and breast-feeding duration (months)) for each cytokine. Estimated marginal means (EMMs) were used for pairwise comparisons between factor levels. Statistical significance was set at *p* < 0.05. BH-adjusted p-values were reported for pairwise comparisons involving HIV status.

#### Overall cytokine correlations

Pairwise Pearson correlation coefficients (r) were calculated using complete observations to generally assess correlations between cytokines measured in plasma and saliva. The resulting correlation matrix was used to generate a heatmap illustrating the direction and magnitude of within- and between-compartment correlations.

#### Overall cytokine profiles

Using the *vegan* package, Permutational multivariate analysis of variance (PERMANOVA) tested differences in multivariate cytokine profiles between groups, while Permutational analysis of multivariate dispersion (PERMDISP) tested for differences in within-group dispersion around group centroids. PERMANOVA p-values were calculated using distances derived from log_2_(x+1)-transformed adjusted cytokine concentrations, adjusting for sex, age (months), height (cm), weight (kg), and breast-feeding duration (months) with 9,999 permutations. PERMANOVA R² values quantified the proportion of distance-matrix variation explained by group membership. Principal components analysis (PCA) was used to visualize variation in the profiles.

#### Multivariable models for individual cytokines

For each cytokine, a multivariable regression model assessed associations with HIV status, CD4⁺ T-cell count (cells/mm³), and caries status, adjusting for sex, age (months), height (cm), weight (kg), and breastfeeding duration (months). Estimated marginal means (EMMs) were used for pairwise comparisons between factor levels. BH-adjusted p-values were reported for pairwise comparisons involving HIV status. P-values for the other predictors were unadjusted.

#### Plasma-saliva concordance

<u>Partial</u> spearman correlation analyses were performed separately for saliva and plasma cytokine concentrations within the HI, HEU, and HUU groups, adjusting for age and sex. Cytokine concentrations were log_2_(x+1)-transformed prior to analysis. Pairwise correlation coefficients (rho) and corresponding p-values were calculated, with multiple comparisons controlled using the Benjamini-Hochberg false discovery rate (FDR) procedure. Associations with FDR-adjusted p-values < 0.05 were considered statistically significant.

*Correlation networks* comprising seven matching cytokines (IFN-γ, IL-1α, IL-4, IL-5, IL-6, IL-8, and MCP-1) between plasma and saliva were constructed separately for each clinical group and biological compartment. Cytokines were represented as nodes, with edges representing statistically significant positive or negative partial Spearman correlations. Positive and negative edges indicated direct and inverse associations, respectively. Network density was evaluated across absolute correlation magnitude thresholds ranging from |ρ| ≥ 0.40 to |ρ| ≥ 0.50, in increments of 0.01, to assess the sensitivity of network connectivity to threshold selection.

*Network density* was calculated as the ratio of observed to maximum possible edges in each undirected network.^61,62^ With seven cytokines, each network comprised a maximum of 21 possible edges (n(n−1)/2). The numbers of total, positive, and negative edges, together with network density, were summarized descriptively across clinical groups, biological compartments, and correlation thresholds.

*For node-level centrality analysis*, networks were constructed using all statistically significant cytokine associations (FDR < 0.05), without imposing an additional minimum correlation coefficient threshold. Edge weights were defined by the absolute magnitude of the partial Spearman correlation coefficients. Network centrality measures included degree (number of direct connections), strength (sum of the absolute weights of connections), betweenness (frequency with which a node lies on the shortest paths between other nodes), closeness, eigenvector centrality (importance of a node based on its connections to other highly connected nodes) and clustering coefficient (extent to which a node’s neighboring nodes are interconnected).^63–65^

#### Effect modification

To explore whether caries-cytokine associations differed by HIV status or immune status, HIV status × caries and CD4⁺ T-cell count × caries interaction terms were added to the cytokine-specific models. Where an interaction was significant, stratum-specific estimates were obtained from estimated marginal means and trends, with BH adjustment.

#### Sensitivity analyses

The multivariate and regression analyses were repeated with maternal education and employment status as additional covariates in the children with these data (n = 400). In the children with salivary flow rate data (n = 203), multiple linear regression assessed associations between salivary flow rate and salivary cytokine concentrations, adjusting for sex, age (months), height (cm), weight (kg), and breastfeeding duration (months).

## Ethical Considerations

The study was approved by the institutional review boards of the University of Maryland Baltimore (HP-00084081), Rutgers, The State University of New Jersey (Pro2019002047), and the University of Benin Teaching Hospital (ADM/E22/A/VOL.VII/14713). Written, informed consent was obtained from all participating parents, guardians, and caregivers of these children before the initiation of the recruitment process. The participating staff in the study thoroughly explained the activities in both English and Pidgin (broken) English, and follow-up questions were given afterward to confirm that the guardians were aware of what the study entails.

## Supporting information

All Supplementary Figures

All Supplementary Tables

## Data Availability

All data produced in the present study are available upon reasonable request to the authors

## Acknowledgments

The authors gratefully acknowledge the families and staff of the DOMHaIN study for their invaluable participation and support. We also extend our appreciation to the University of Benin Teaching Hospital and the Institute of Human Virology Nigeria for their collaboration and assistance in facilitating this research. This work was supported by the National Institutes of Health/National Institute of Dental and Craniofacial Research (R01DE028154).

