## Supplementary material for "Distinct Systemic and Salivary Cytokine Profiles in Children with Perinatal HIV Exposure and Infection": All Supplementary Figures

*Figure 1a*:

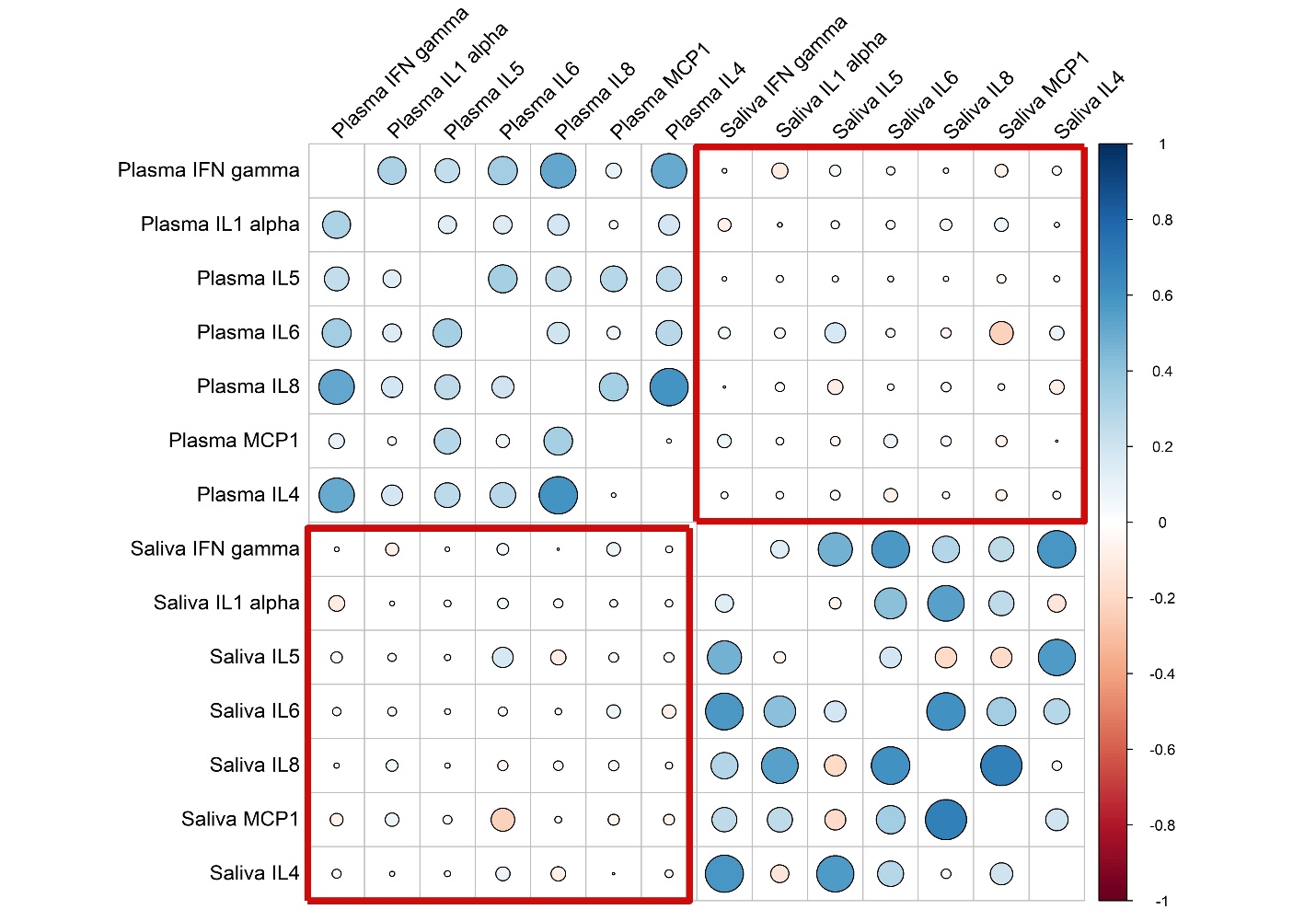

***Figure 1a: Correlations among and between plasma and salivary cytokines among children living with HIV (HI)****. Heatmap illustrating the unadjusted pairwise Pearson correlations (r) among cytokines measured in plasma and saliva. Correlation coefficients are represented by circle size and color, with larger circles indicating stronger correlations. Blue indicates positive correlations, while red indicates negative correlations. The red outlined regions highlight between-compartment cytokine correlations, while the remaining regions represent within-compartment correlations among plasma and salivary cytokines.*

*Figure 1b*:

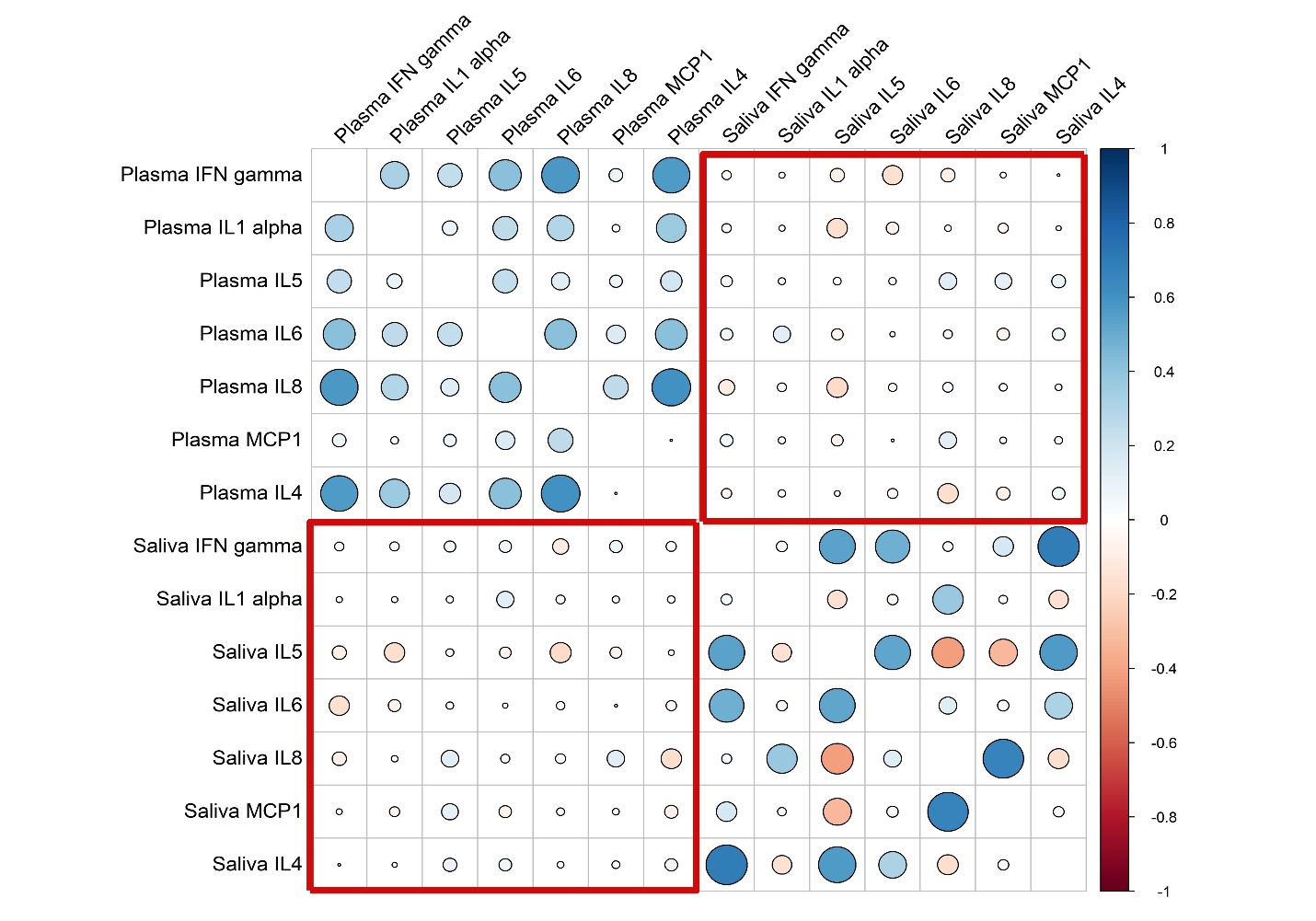
***Figure 1b: Correlations among and between plasma and salivary cytokines among children who were HIV-exposed but uninfected (HEU)****. Heatmap illustrating the unadjusted pairwise Pearson correlations (r) among cytokines measured in plasma and saliva. Correlation coefficients are represented by circle size and color, with larger circles indicating stronger correlations. Blue indicates positive correlations, while red indicates negative correlations. The red outlined regions highlight between-compartment cytokine correlations, while the remaining regions represent within-compartment correlations among plasma and salivary cytokines.*

*Figure 1c*:

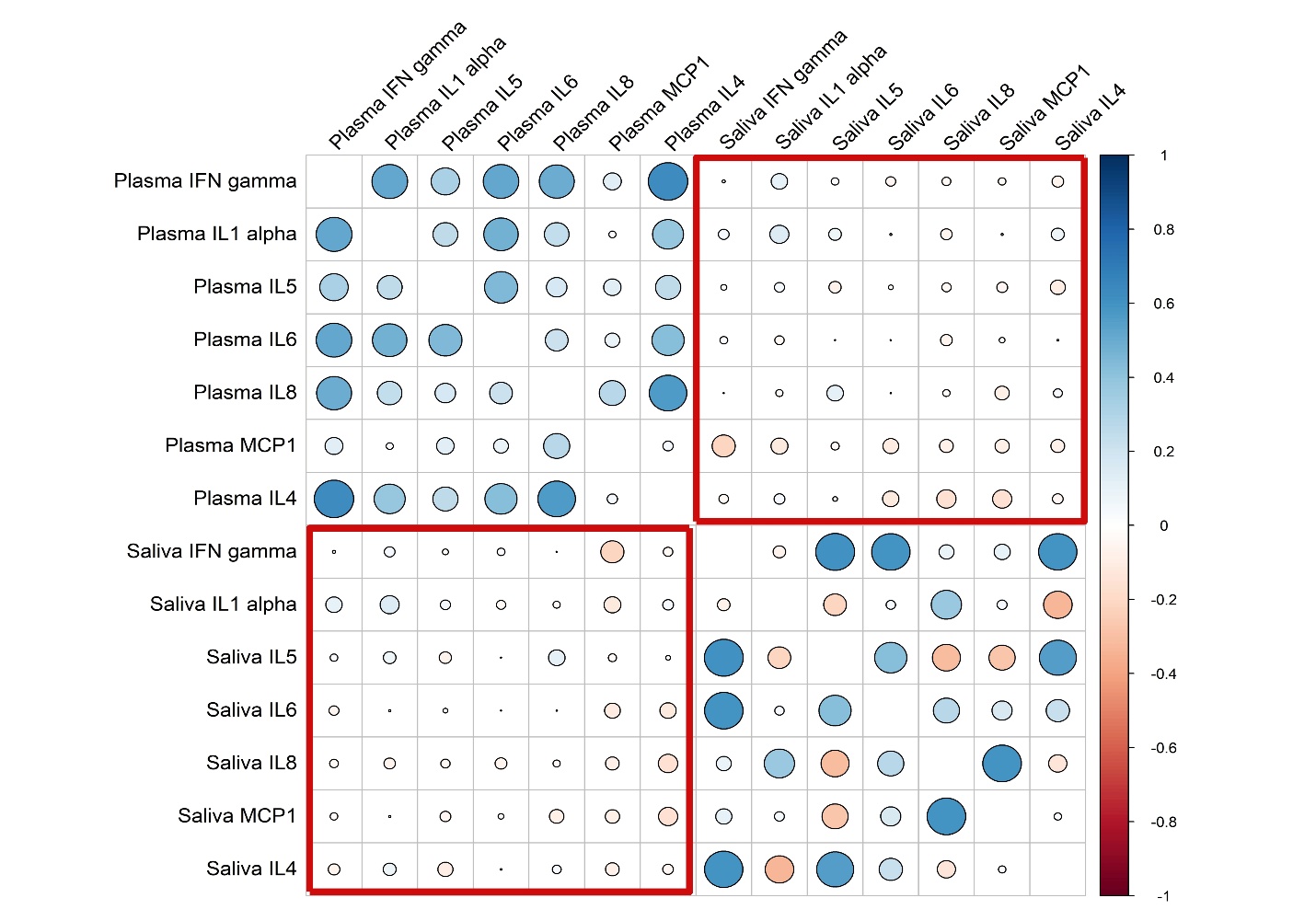
***Figure 1c: Correlations among and between plasma and salivary cytokines among children who were HIV-unexposed and uninfected (HUU)****. Heatmap illustrating the unadjusted pairwise Pearson correlations (r) among cytokines measured in plasma and saliva. Correlation coefficients are represented by circle size and color, with larger circles indicating stronger correlations. Blue indicates positive correlations, while red indicates negative correlations. The red outlined regions highlight between-compartment cytokine correlations, while the remaining regions represent within-compartment correlations among plasma and salivary cytokines.*
