## Supplementary material for "Distinct Systemic and Salivary Cytokine Profiles in Children with Perinatal HIV Exposure and Infection": All Supplementary Tables

**Supplementary Tables & Figures**:

*Supplementary Table 1*:

| Principal Component 1 | | Principal Component 2 | |
| --- | --- | --- | --- |
| Cytokines | Loadings | Cytokines | Loadings |
| IFN-γ | 0.421 | IL-1β | 0.297 |
| IL-4 | 0.375 | IL-1α | 0.292 |
| IL-1β | 0.374 | IL-4 | 0.265 |
| IL-8 | 0.363 | IFN-γ | 0.143 |
| IL-6 | 0.342 | IL-8 | 0.113 |
| IL-1α | 0.324 | IL-6 | -0.206 |
| IL-5 | 0.267 | IL-5 | -0.323 |
| IL-10 | 0.263 | TNF-α | -0.366 |
| TNF-α | 0.174 | IL-10 | -0.458 |
| MCP-1 | 0.136 | MCP-1 | -0.482 |

***Supplementary Table 1: Plasma cytokine loadings on PC1 and PC2.*** *Plasma cytokines are ranked from highest to lowest according to their loadings on PC1 and PC2. Higher loading values indicate a greater contribution to the corresponding principal component. All 10 cytokines had positive loadings on PC1, whereas only IL-1β, IL-1α, IL-4, IFN-γ, and IL-8 had positive loadings on PC2.*

*Supplementary Table 2*:

| Principal Component 1 | | Principal Component 2 | |
| --- | --- | --- | --- |
| Cytokines | Loadings | Cytokines | Loadings |
| IFN-γ | 0.563 | IL-5 | 0.373 |
| IL-4 | 0.477 | IL-4 | 0.220 |
| IL-6 | 0.467 | IFN- γ | 0.013 |
| IL-5 | 0.429 | IL-6 | -0.181 |
| MCP-1 | 0.175 | IL-1α | -0.395 |
| IL-8 | 0.151 | MCP-1 | -0.496 |
| IL-1α | 0.020 | IL-8 | -0.614 |

***Supplementary Table 2: Saliva cytokine loadings on PC1 and PC2.*** *Saliva cytokines are ranked from highest to lowest according to their loadings on PC1 and PC2. Higher loading values indicate a greater contribution to the corresponding principal component. All 7 cytokines had positive loadings on PC1, whereas only IL-5, IL-4, and IFN-γ had positive loadings on PC2.*

*Supplementary Table 3*: Supplementary Table 3 is given as a excel file.

*Supplementary Table 4*:

| **Cytokine** | **Predictor/Comparison** | **Estimate** | **Std. error** | **p-value** | **Partial R^2^** |
| --- | --- | --- | --- | --- | --- |
| IL-8 | **Salivary Flow Rate (mL/min)**: | | | | |
|  | Per 1-mL/min increase | -1.615 | 0.581 | 0.00597 | 0.039 |
| MCP-1 | **Salivary Flow Rate (mL/min)**: | | | | |
|  | Per 1-mL/min increase | -1.723 | 0.458 | 0.000224 | 0.069 |

***Supplementary Table 4. Associations between salivary flow rate and salivary cytokine concentrations.*** *Linear regression estimates, standard errors, p-values, and partial R² values are presented for the associations between salivary flow rate (mL/min) and salivary IL-8 and MCP-1 concentrations. Estimates represent the change in cytokine concentration associated with each 1-mL/min increase in salivary flow rate. Partial R² values indicate the proportion of variance uniquely explained by salivary flow rate in each model.*

***Supplementary Table 5a***:

| **Cytokine** | **Model term** | **β coefficient** | ***p*-value** | **Interpretation** |
| --- | --- | --- | --- | --- |
| IL-1α | **HI** | **+1.251** | **0.022** | **Higher IL-1α in HI vs. reference HIV group among children with active caries** |
|  | HUU | +1.156 | 0.050 | No statistically significant difference vs. reference HIV group among children with active caries. But approaching significance. |
|  | **Caries-free** | **+1.253** | **0.008** | **Higher IL-1α in caries-free vs. active-caries children in the reference HIV group** |
|  | **HI × caries-free** | **−1.798** | **0.003** | **Significant interaction between HI status and caries status** |
|  | **HUU × caries-free** | **−1.370** | **0.027** | **Significant interaction between HUU status and caries status** |
| IL-1β | HI | +0.439 | 0.320 | No statistically significant difference vs. reference HIV group among children with active caries |
|  | HUU | +0.319 | 0.503 | No statistically significant difference vs. reference HIV group among children with active caries |
|  | Caries-free | +0.728 | 0.055 | No statistically significant caries-status association in the reference HIV group. But approaching significance. |
|  | **HI × caries-free** | **−0.971** | **0.047** | **Significant interaction between HI status and caries status** |
|  | HUU × caries-free | −0.747 | 0.136 | No significant interaction |

***Supplementary Figure 5a. Associations between HIV status, dental caries status, and plasma IL-1α and IL-1β concentrations.*** *Multivariable linear regression models evaluated associations of HIV status (HI and HUU, relative to HEU), dental caries status (caries-free relative to active caries), and their interactions with cytokine concentrations. β coefficients represent estimated differences in log_2_(x+1)-transformed cytokine concentrations relative to the reference categories. Statistical significance was defined as p<0.05.*

***Supplementary Table 5b***:

| **Cytokine** | **HIV exposure/infection group** | **Caries comparison** | **β estimate** | **BH-adjusted p-value** |
| --- | --- | --- | --- | --- |
| IL-1α | **HEU** | **Active caries vs. caries-free** | **−1.253** | **0.008** |
|  | HI | Active caries vs. caries-free | +0.545 | 0.148 |
|  | HUU | Active caries vs. caries-free | +0.117 | 0.772 |
| IL-1β | HEU | Active caries vs. caries-free | −0.728 | 0.055 |
|  | HI | Active caries vs. caries-free | +0.243 | 0.424 |
|  | HUU | Active caries vs. caries-free | +0.019 | 0.954 |

***Supplementary Figure 5b. Associations between dental caries status and plasma IL-1α and IL-1β concentrations* stratified by *HIV exposure/infection groups.*** *Comparisons of cytokine concentrations between children with active caries and those who were caries-free were performed separately among children living with HIV (HI), HIV-exposed uninfected (HEU), and HIV-unexposed uninfected (HUU) children. β estimates represent differences in log_2_(x+1)-transformed cytokine concentrations between children with active caries and those who were caries-free. Positive estimates indicate higher concentrations in children with active caries, whereas negative estimates indicate higher concentrations in caries-free children. P-values were adjusted for multiple comparisons using the Benjamini–Hochberg (BH) procedure. Statistical significance was defined as BH-adjusted p<0.05.*

***Supplementary Table 5c***:

| **Cytokine** | **Caries status** | **HIV-group comparison** | **β estimate** | **BH-adjusted p-value** |
| --- | --- | --- | --- | --- |
| IL-1α | Active caries | HI vs. HEU | +1.252 | 0.067 |
|  |  | HUU vs. HEU | +1.156 | 0.076 |
|  |  | HI vs. HUU | +0.095 | 0.845 |
|  | Caries-free | HI vs. HEU | -0.546 | 0.219 |
|  |  | HUU vs. HEU | -0.214 | 0.487 |
|  |  | HI vs. HUU | -0.333 | 0.417 |
| IL-1β | Active caries | HI vs. HEU | +0.439 | 0.755 |
|  |  | HUU vs. HEU | +0.319 | 0.755 |
|  |  | HI vs. HUU | +0.120 | 0.760 |
|  | Caries-free | HI vs. HEU | -0.532 | 0.093 |
|  |  | HUU vs. HEU | -0.427 | 0.129 |
|  |  | HI vs. HUU | -0.104 | 0.674 |

***Supplementary Figure 5c. Comparisons of plasma IL-1α and IL-1β concentrations across HIV exposure/infection groups stratified by dental caries status.*** *Pairwise comparisons of cytokine concentrations were performed between children living with HIV (HI), HIV-exposed uninfected (HEU), and HIV-unexposed uninfected (HUU) children, separately among children with active caries and those who were caries-free. β estimates represent differences in log_2_(x+1)-transformed cytokine concentrations between HIV exposure/infection groups, with positive estimates indicating higher concentrations in the first group listed. P-values were adjusted for multiple comparisons using the Benjamini–Hochberg (BH) procedure. Statistical significance was defined as BH-adjusted p<0.05.*

***Supplementary Table 5d***:

| **Cytokine** | **Model term** | **β coefficient** | ***p*-value** | **Interpretation** |
| --- | --- | --- | --- | --- |
| IL-1α | **CD4^+^ count** | **−0.001** | **0.007** | **Higher CD4^+^ counts associated with lower IL-1α among children with active caries** |
|  | **Caries-free** | **−1.221** | **0.028** | **Lower IL-1α in caries-free children at CD4^+^ count = 0** |
|  | **CD4^+^ count × caries-free** | **+0.001** | **0.011** | **Significant interaction between CD4^+^ count and caries status** |
|  | HI | **−**0.310 | 0.264 | No statistically significant difference vs. reference HIV group, adjusted for other variables |
|  | HUU | **−**0.055 | 0.849 | No statistically significant difference vs. reference HIV group, adjusted for other variables |
| IFN-γ | CD4^+^ count | −4.572 x 10^-4^ | 0.142 | Higher CD4^+^ counts were not statistically associated with lower IFN-γ among children with active caries |
|  | **Caries-free** | **−9.172 x 10^-1^** | **0.009** | **Lower IFN-γ in caries-free children at CD4^+^ count = 0** |
|  | **CD4^+^ count × caries-free** | **+7.567 x 10^-4^** | **0.029** | **Significant interaction between CD4^+^ count and caries status** |
|  | HI | **−**2.664 x 10^-1^ | 0.131 | No statistically significant difference vs. reference HIV group, adjusted for other variables |
|  | **HUU** | **−3.692 x 10^-1^** | **0.045** | **Lower IFN-γ in HUU vs. HEU, adjusted for other variables** |

***Supplementary Figure 5d. Associations of CD4^+^ count, dental caries status, and HIV exposure/infection group with plasma IL-1α and IFN-γ concentrations.*** *Multivariable linear regression models evaluated associations of CD4^+^ count, dental caries status, and HIV exposure/infection group with cytokine concentrations, including an interaction term between CD4^+^ count and caries status. CD4^+^ count was modeled as a continuous variable, and caries-free status was compared with active caries as the reference group. HI and HUU groups were compared with HEU as the reference group. β coefficients represent estimated changes in log_2_(x+1)-transformed cytokine concentrations associated with each model term. Significant CD4^+^ count-by-caries status interactions indicate that the association between CD4^+^ count and cytokine concentrations differed according to caries status. Statistical significance was defined as p<0.05.*

***Supplementary Table 5e***:

| **Cytokine** | **Caries status** | **β (CD4 slope)** | **SE** | **95% CI** | **p-value** |
| --- | --- | --- | --- | --- | --- |
| IL-1α | **Active caries** | **-1.33 x 10^-3^** | **4.90 x 10^-4^** | **-2.30e-03 - -3.70e-03** | **0.007** |
|  | Caries-free | 6.07 x 10^-5^ | 2.60 x 10^-4^ | -4.50e-04 – 5.72e-04 | 0.815 |
| IFN-γ | Active caries | -4.57 x 10^-4^ | 3.11 x 10^-4^ | -1.07e-03 – 1.54e-04 | 0.142 |
|  | Caries-free | 3.00 x 10^-4^ | 1.65 x 10^-4^ | -2.44e-05 – 6.24e-04 | 0.070 |

***Supplementary Figure 5e. Associations between CD4^+^ count and plasma IL-1α and IFN-γ concentrations stratified by dental caries status.*** *CD4^+^ count–cytokine associations were evaluated separately among children with active caries and those who were caries-free using multivariable linear regression models. β coefficients represent the estimated change in log_2_(x+1)-transformed cytokine concentrations per unit increase in CD4^+^ count. Standard errors (SE) and 95% confidence intervals (CI) are provided for each estimated slope. Models were adjusted for HIV exposure/infection group and other covariates. Statistical significance was defined as p<0.05.*

*Supplementary Table 6*:

| **Cytokine** | **QC Pairs** | **Mean CV%** | **Median CV%** | **CV% Range (Q1 – Q3)** | **% CV <20%** |
| --- | --- | --- | --- | --- | --- |
| IFN-γ | 57 | 7.59 | 4.10 | 1.6 – 7.9 | 91.2 |
| IL-1α | 57 | 11.08 | 6.60 | 2.9 – 14.1 | 80.7 |
| IL-1β | 70 | 6.13 | 3.90 | 2.2 – 9.8 | 98.6 |
| IL-2 | 75 | 7.78 | 4.79 | 1.8 – 9.7 | 89.3 |
| IL-4 | 77 | 8.66 | 5.30 | 2.3 – 12.4 | 93.5 |
| IL-5 | 78 | 8.47 | 4.65 | 1.75 – 9.9 | 89.7 |
| IL-6 | 72 | 9.13 | 5.60 | 2.7 – 13.9 | 88.9 |
| IL-8 | 74 | 8.38 | 6.05 | 2.53 – 10.6 | 90.5 |
| IL-10 | 74 | 7.10 | 4.80 | 2.33 – 10.83 | 94.6 |
| MCP-1 | 71 | 9.11 | 5.50 | 2.7 – 11.7 | 93.0 |
| TNF-α | 78 | 9.75 | 5.45 | 3.22 – 10.3 | 87.2 |

***Supplementary Table 6. Plasma Intra-assay Coefficient of Variation.*** *Intra-assay coefficient of variation is calculated for each cytokine between duplicate observations (QC pairs) within assay runs in plasma. Mean CV%, Median CV%, and CV% Interquartile Range are reported. The percentage of intra-assay coefficients of variation less than 20% are also reported.*

*Supplementary Table 7*:

| **Cytokine** | **QC Pairs** | **Mean CV%** | **Median CV%** | **CV% Range (Q1 – Q3)** | **% CV <20%** |
| --- | --- | --- | --- | --- | --- |
| IFN-γ | 63 | 8.43 | 6.40 | 2.85 – 10.4 | 88.9 |
| IL-1α | 67 | 10.44 | 8.10 | 4.00 – 14.05 | 86.6 |
| IL-1β | 74 | 8.86 | 5.65 | 3.92 – 11.58 | 91.9 |
| IL-2 | 74 | 7.85 | 6.05 | 2.55 – 9.83 | 94.6 |
| IL-4 | 74 | 8.87 | 7.15 | 3.30 – 12.28 | 87.8 |
| IL-5 | 79 | 6.95 | 4.90 | 2.20 – 9.10 | 94.9 |
| IL-6 | 76 | 7.80 | 5.90 | 2.50 – 11.75 | 93.4 |
| IL-8 | 78 | 6.91 | 5.30 | 1.90 – 10.88 | 96.2 |
| IL-10 | 79 | 5.56 | 3.60 | 1.50 – 6.95 | 97.5 |
| MCP-1 | 67 | 8.56 | 6.00 | 2.95 – 9.95 | 94.0 |
| TNF-α | 72 | 10.10 | 7.00 | 4.00 – 11.95 | 88.9 |

***Supplementary Table 7. Saliva Intra-assay Coefficient of Variation.*** *Intra-assay coefficient of variation is calculated for each cytokine between duplicate observations (QC pairs) within assay runs in saliva. Mean CV%, Median CV%, and CV% Interquartile Range are reported. The percentage of intra-assay coefficients of variation less than 20% are also reported.*

*Supplementary Table 8*:

| **a) Plasma Inter-Assay CV%** | | | **b) Saliva Inter-Assay CV%** | | |
| --- | --- | --- | --- | --- | --- |
| **Cytokine** | **Mean Inter-Run CV%** | **Batch-adjusted Inter-Run CV%** | **Cytokine** | **Mean Inter-Run CV%** | **Batch-adjusted Inter-Run CV%** |
| IFN-γ | 80.91 | 14.65 | IFN-γ | 15.37 | 14.65 |
| IL-1α | 85.69 | 27.82 | IL-1α | 16.22 | 27.82 |
| IL-1β | 16.90 | 17.14 | IL-1β | 30.12 | 17.14 |
| IL-2 | 78.52 | 4.09 | IL-2 | 14.69 | 4.09 |
| IL-4 | 78.37 | 3.46 | IL-4 | 25.41 | 3.46 |
| IL-5 | 78.56 | 3.15 | IL-5 | 30.00 | 3.15 |
| IL-6 | 77.64 | 2.76 | IL-6 | 30.04 | 2.76 |
| IL-8 | 86.60 | 4.11 | IL-8 | 29.87 | 4.11 |
| IL-10 | 80.71 | 21.95 | IL-10 | 31.63 | 21.95 |
| MCP-1 | 68.52 | 25.2 | MCP-1 | 27.53 | 25.2 |
| TNF-α | 78.94 | 30.19 | TNF-α | 31.23 | 30.19 |

***Supplementary Table 8. Inter-assay Coefficient of Variation.*** *Mean inter-assay coefficient of variation is calculated for each cytokine between assay runs in a) plasma and b) saliva. Batch-adjusted inter-run CV% were calculated using ComBat.*

*Supplementary Table 9*:

| **Compartment** | **Cytokine** | **LLOD (pg/mL)** | **Below LLOD, n(%)** | **At or above LLOD, n(%)** | **>30% below LLOD** |
| --- | --- | --- | --- | --- | --- |
| a) Plasma Cytokines | IFN-γ | 1.42 | 77 (16.5%) | 390 (83.5%) | No |
|  | IL-1α | 3.48 | 134 (28.7%) | 333 (71.3%) | No |
|  | IL-1β | 0.79 | 97 (20.8%) | 370 (79.2%) | No |
|  | IL-2 | 0.46 | 178 (38.1%) | 289 (61.9%) | Yes |
|  | IL-4 | 0.29 | 99 (21.2%) | 368 (78.8%) | No |
|  | IL-5 | 0.21 | 34 (7.3%) | 433 (92.7%) | No |
|  | IL-6 | 0.20 | 111 (23.8%) | 356 (76.2%) | No |
|  | IL-8 | 0.58 | 130 (27.8%) | 337 (72.2%) | No |
|  | IL-10 | 1.76 | 31 (6.6%) | 436 (93.4%) | No |
|  | MCP-1 | 3.24 | 1 (0.2%) | 466 (99.8%) | No |
|  | TNF-α | 5.75 | 4 (0.9%) | 463 (99.1%) | No |
| b) Saliva Cytokines | IFN-γ | 1.42 | 103 (22.1%) | 364 (77.9%) | No |
|  | IL-1α | 3.48 | 82 (17.6%) | 385 (82.4%) | No |
|  | IL-1β | 0.79 | 212 (45.4%) | 255 (54.6%) | Yes |
|  | IL-2 | 0.46 | 306 (65.5%) | 161 (34.5%) | Yes |
|  | IL-4 | 0.29 | 72 (15.4%) | 395 (84.6%) | No |
|  | IL-5 | 0.21 | 116 (24.8%) | 351 (75.2%) | No |
|  | IL-6 | 0.20 | 24 (5.1%) | 443 (94.9%) | No |
|  | IL-8 | 0.58 | 3 (0.6%) | 464 (99.4%) | No |
|  | IL-10 | 1.76 | 167 (35.8%) | 300 (64.2%) | Yes |
|  | MCP-1 | 3.24 | 3 (0.6%) | 464 (99.4%) | No |
|  | TNF-α | 5.75 | 307 (65.7%) | 160 (34.3%) | Yes |

***Supplementary Table 9. Cytokine detectability in plasma and saliva.*** *The table summarizes the lower limit of detection (LLOD) for each cytokine and the number and percentage of observations below or at/above the LLOD in each biological compartment. Missing or unreported measurements were classified as below the LLOD. Cytokines with more than 30% of observations below the LLOD were flagged.*
